# Vitamin D deficiency and depressive episodes in adolescents and young adults: systematic review and meta-analysis

**DOI:** 10.1101/2025.02.11.25321917

**Authors:** Joseph P. Nano, William A. Catterall, Violet Filer, Salar Khaleghzadegan, Ricardo Almazan, Heather B. Blunt, Renata W. Yen

## Abstract

**Background:** We conducted a systematic review and meta-analysis to examine the association between vitamin D deficiency and depression in adolescents (10–17 years) and young adults (18–39 years).

**Methods:** We searched databases and reference lists (inception to October 2025, limited to English studies). We extracted data and assessed study quality using the Newcastle-Ottawa Scale. We used random-effects meta-analyses to determine mean difference scores in vitamin D serum levels (depressed vs. non-depressed), odds ratios for vitamin D categories (insufficiency and deficiency), and subgroup analyses (gender and age). We assessed statistical heterogeneity using I^2^, with fixed-effect modeling as a sensitivity analysis.

**Results:** From 11,309 citations, we found 20 studies between 2010 and 2025 (mean=27,956; range: 51 to 483,683). Twelve studies (60%) found that vitamin D insufficiency (50–75 nmol/L) and deficiency (<50 nmol/L) were significantly associated with depression. Our meta-analysis revealed that the depressed population of young adults and adolescents had lower mean vitamin D levels than non-depressed participants (random-effect MD=-17.77 nmol/L [-30.12,-5.42]; p<0.001; I²=95%). We found a significant association between depression and vitamin D deficiency (random-effect OR=2.06 [1.02, 4.17], p<0.001; I^2^=95%) but no association with vitamin D insufficiency (random-effect OR=0.94 [0.74, 1.21]; I^2^=0%). Fixed-effect and random-effects models produced consistent results. Female participants had a stronger association (random-effect OR=1.88 [1.29, 2.74], I^2^=97%) and no significant association among those aged 25 to 39 years old (random-effect OR=1.14 [0.78, 1.65], p<0.001, I^2^=94%).

**Conclusion:** Our systematic review suggested that vitamin D deficiency is significantly associated with depression, with important variations by sex and age.

## INTRODUCTION

According to the World Health Organization, 4.4% of the world’s population suffers from depression.^1^ Depression is a mood disorder that can lead to changes in sleep patterns, appetite, daily energy, and feelings.^2–4^ Depression typically starts in adolescence and may continue into adulthood.^5^ There are different types of depression: major depression, persistent depressive disorder, perinatal depression, seasonal affective disorder, and depression with symptoms of psychosis.^2^ The causes of depression are multifactorial, including biological, genetic, environmental, and psychosocial causes.^1^ An estimated 4.1 million adolescents in the United States (17% of adolescents in the U.S. population) had at least one major depressive episode in 2015 and according to 2023 Gallup polls 34% of U.S. young adults reported having depression during their lifetime.^1,6^ Thus, depression places an enormous health burden on adolescents and young adults.^7^ Researchers hypothesize that major depression is projected to be one of the three leading disease burdens affecting adolescents and adults by 2030.^8^ Furthermore, depression may affect not only adolescents and young adults but also parents, families, schools, healthcare providers, pharmaceutical companies, public policymakers, and researchers. It is essential to find solutions to preventing and reducing symptoms of depression.^9^

Vitamin D is a neurosteroid hormone essential for neuroprotection and is suggested to play a critical role in the regulation of mood behaviors because its receptors are widely spread throughout the human brain.^10,11^ The total serum 25-hydroxyvitamin D (25(OH)D) is the primary circulating form of vitamin D and considered the best indicator of vitamin D level within the human body.^12^ Vitamin D serum levels in the body are influenced by environmental factors, such as exposure to sunlight, which stimulates cutaneous synthesis of vitamin D in the skin, as well as dietary intake, and vitamin D supplementation.^13,14^ Vitamin D is an essential nutrient that plays a critical role in several physiological processes, including bone health, immune regulation, and cardiometabolic function, and has been widely studied in relation to various health outcomes.^15–18^ Vitamin D status is also increasingly studied in its relationship with depression in population-based studies.^19^ Recent data from a large-scale genome-wide association study that focused on populations of European ancestry did not show a causal relationship between vitamin D deficiency and depression.^19^

There is still a controversy on whether vitamin D deficiency is associated with depression. While some studies have shown a strong association between Vitamin D and depression,^20–23^ others have demonstrated no association.^24–27^ This variability may be explained by differences in study populations, geographic variation in sunlight exposure that influences vitamin D status, and differences in underlying health conditions across study samples.^13^ Some reviews have examined the association between vitamin D and depression, primarily during adulthood (e.g., aged 18–59 years) or elderly populations, as well as in individuals with medical comorbidities such as cardiovascular disease, diabetes, or chronic inflammatory conditions.^28^ While these studies have provided important insights into the potential association between vitamin D deficiency and depressive symptoms during a broad range of adulthood, the evidence remains limited for specifically younger populations (aged 18–39 years). In particular, there is a clear gap in the literature regarding the relationship between vitamin D serum levels and depression among adolescents and young adults, which is a critical developmental period when mood disorders often first emerge.^29,30^ Furthermore, adolescents and young adults are more likely to be considered “healthy individuals” compared to elderly populations, which may influence the relationship between vitamin D and depression. As such, studying healthy individuals minimizes the possibility that observed associations between vitamin D and depression are explained by other health conditions and behaviors.

Some studies have investigated the association between vitamin D supplementation and depression.^27^ It is important to distinguish between studies examining vitamin D serum levels and those evaluating vitamin D supplementation. Observational studies assess vitamin D status as an exposure associated with health outcomes, whereas supplementation studies evaluate the effects of increasing vitamin D intake through interventions. While interventional studies focus on supplementation effects on mental health outcomes, our review complements this literature by examining the association between vitamin D serum levels and depression specifically among adolescents and young adults. Establishing observational associations between vitamin D status and depression may provide an important foundational framework for interpreting and informing future supplementation trials in this population. This is the first systematic review to focus on the combined population of adolescents (10–17 years) and young adults (18–39 years) for this topic. The purpose of this study is to determine if there is an association between vitamin D serum levels and depression among adolescents and young adults.

## METHODS

### Eligibility criteria

#### I. Study design

We followed standard analysis procedures outlined by the Cochrane Handbook and the PRISMA guidelines when reporting methods and results for this systematic review and meta-analysis.^31,32^ We created a protocol with Open Science Framework (OSF) in February 2023 to guide the inclusion of studies. We registered our protocol in November 2023 (https://osf.io/ujrt4) (See **Appendix 7** for systematic review protocol modifications).

#### II. Inclusion criteria

For exposure, we considered studies examining vitamin D serum levels categorized as deficient (<50 nmol/L) and insufficient (50–75 nmol/L), with a control group defined by vitamin D sufficiency (>75 nmol/L).

Our population of interest included adolescents and young adults. We defined adolescents as individuals aged 10 to 17 years based on the World Health Organization definition of adolescence.^33^ We defined young adults as individuals aged 18 to 39 years, consistent with age classifications frequently used in epidemiological and population health research examining early adulthood (see our inclusion criteria table in **Appendix 1**).^77,78^ This is the most relevant population to our research topic, because depression is more likely to start during adolescence. Also, choosing the population age of young adulthood (18 to 39) minimizes confounding from age-related chronic diseases because this population is less likely to experience morbidity that would increase the risk for depression.^79^ We considered studies that included broader age ranges (e.g., 18 to 75 years) eligible if they provided subgroup analyses specific to adolescents or young adults that could be extracted. When necessary, we contacted study investigators to obtain age-specific data, if available.

The primary outcome of interest was depression, assessed using validated instruments. We excluded studies that did not match our language or age criteria. Further information on the specific inclusion criteria and outcomes of interest can be found in **Appendix 1**, and the PubMed search terms used are listed in **Appendix 2**.

We included studies regardless of geographic location. Country of origin was recorded to provide contextual information related to differences in sunlight exposure, dietary vitamin D intake, and population characteristics.

### Data sources

Our final search covers studies from inception to October 2025. Our initial search included all dates up to the date we ran the search, July 20, 2023. After two years, we updated the search and screened references from July 1st, 2023 up to October 30, 2025. We used exploded MeSH terms and keywords to generate themes related to vitamin D serum levels and depression. We used the boolean operator “AND” to find the intersection between themes (See **Appendix 3** for search strategies). We performed electronic searches in MEDLINE (via PubMed), The Cochrane Library, Cumulated Index in Nursing and Allied Health Literature (CINAHL), PsycINFO, Scopus, reference reviews through Google Scholar, and ClinicalTrials.gov.^34–37^ In collaboration with a reference librarian (HBB), we (JN and VF) developed a list of keywords and subject headings in PubMed MEDLINE and ran it in each database (See **Appendix 2** for PubMed search terms table). We also reviewed the reference lists of our included studies. For all studies included that pulled from the same dataset, we ensured the data used was not from overlapping time periods or overlapping participants.

### Selection process

Two independent reviewers (JN and WC) used Rayyan to screen studies from inception to October 2025 in two stages: first title and abstract screening followed by full-text screening.^38^ We removed studies that did not meet inclusion criteria. During title and abstract screening, the two reviewers agreed on 92.7% of records. Then, two independent and blinded reviewers (JN and WC) conducted an assessment of the full text of each study based on inclusion criteria. The reviewers resolved data discrepancies after review. Inter-rater agreement during the full-text screening and study selection process was high, with approximately 92.0% agreement between reviewers (Weighted kappa = 0.956).

### Data collection process

Data extraction tables were developed a priori based on the *Cochrane Handbook for Systematic Reviews of Interventions* (version 6.3, Chapters 5 and 10). Two reviewers (JN and WC) independently extracted data into three standardized tables capturing study characteristics, exposure and outcome variables, and risk of bias assessments. Extracted data included study design, population characteristics, vitamin D measurement methods, depression assessment tools, effect estimates, and covariates. The reviewers (JN and WC) used a piloted standardized data collection form and performed independent double data extraction (See **Appendix 4** for a blank data collection form). Reviewers extracted information about the outcome, authors, publication year, country, type of study design, aim(s) and research questions, sample size, follow-up, and control condition.

When studies used non-standard cutoffs or reported vitamin D as a continuous variable, values were reclassified according to the Institute of Medicine (IOM) thresholds and the Endocrine Society (ES) guidelines (deficient <50 nmol/L, insufficient 50–75 nmol/L, sufficient >75 nmol/L) when data permitted.^39,40^ When studies used non-standard cutoffs or reported vitamin D as a continuous variable, values were reclassified according to commonly used clinical thresholds for serum 25-hydroxyvitamin D (deficient <50 nmol/L, insufficient 50–75 nmol/L, sufficient >75 nmol/L) when data permitted. If a study reported additional categories, such as severe deficiency, we only selected categories whose serum vitamin D range most closely aligned with the IOM and ES cut-offs for inclusion in the meta-analysis.

For example, when vitamin D concentrations were reported in ng/mL, we converted them to nmol/L using the standard conversion factor (1 ng/mL = 2.5 nmol/L), in accordance with the Institute of Medicine and Cochrane Handbook recommendations. We extracted adjusted odds ratios and corresponding 95% confidence intervals, when available, from each study to conduct meta-analyses. If multiple adjusted models were reported, we extracted from the most fully adjusted model to account for potential confounding factors. We reported and summarized key covariates included in the adjustment models for each study in **Appendix 9**. For studies that reported effect estimates other than odds ratios (e.g., risk ratios, hazard ratios, mean differences, or correlation coefficients), we applied standard statistical transformations to convert them to log(OR) and corresponding standard errors, following methods described by Borenstein et al. (2011).^41^

### Outcomes

Our primary outcome was depression, which was measured by a validated depression scale or diagnostic tools, including the Hamilton Scale of Depression, Beck Depression Inventory, Depression Diagnostic Criteria, Patient Health Questionnaire, Diagnostic Interview Schedule, Children’s Depression Inventory, and Center for Epidemiologic Studies Depression (**Appendix 5).**^42–48^

We selected secondary outcomes related to the harms and benefits of vitamin D exposure: alcohol consumption, body mass index (BMI), physical activity recreation status, and smoking status. Secondary outcomes were identified prior to the screening process based on theoretical and empirical links with vitamin D status. Studies were not required to report secondary outcomes for inclusion.

### Synthesis methods

For studies that did not provide sufficient quantitative data for meta-analysis, we conducted a qualitative narrative synthesis. For example, we summarized study characteristics (e.g., sample demographics, study design, setting, depression measurement tools, and vitamin D assessment methods). Then, we compared the direction and magnitude of associations reported across studies, noting consistencies, discrepancies, and methodological differences that could explain variation in findings (e.g., geographic region, adjustment for confounders)

We performed a meta-analysis in RevMan (Review Manager version 5.4.1) that examined the association between vitamin D serum levels and depression in two ways. First, we calculated the pooled mean difference (MD) by extracting mean vitamin D serum levels (nmol/L), sample size, and standard deviation of participants who are depressed compared to those who are not depressed for studies that provided this data.^49^ For mean difference analyses, statistical significance was observed when the confidence interval did not cross the null value of 0. Second, we extracted odds ratios, 95% confidence intervals, standard errors to investigate the association between being depressed and different vitamin D categories (vitamin D deficiency and vitamin D insufficiency). For odds ratios, pooled estimates were considered statistically significant when the confidence interval did not cross the null value of 1. When the confidence interval crossed the value of 1, we interpreted the results as not statistically significant (p ≥ 0.05). We assessed heterogeneity using the X^2^ test and I^2^ test.^49^ A p-value threshold of <0.10 on the X^2^ was used to determine statistical significance. We considered a threshold of I^2^ > 50% to indicate significant heterogeneity.^49^ We conducted two types of meta-analyses based on data available: (1) continuous data for overall mean serum levels and (2) dichotomous data (using odds ratio) for two types of vitamin D levels (insufficiency level and deficiency level).

### Assessment of Methodological Quality

We used the Newcastle-Ottawa Quality Assessment Scale to assess the quality of observational studies included in this systematic review.^50,51^ Since the majority of the included studies were cross-sectional, we applied an adapted version of the Newcastle-Ottawa Scale for this study design.^52^ Two independent blinded reviewers (JN and WC) assessed all studies based on three categories, receiving a certain number of stars with a maximum number of 9 stars indicating the highest quality. The three categories were selection, comparability, and outcome (for cohort studies) or exposure (for case-control studies). For example, high quality studies received 7 to 9 stars, moderate quality studies received 4 to 6 stars, and low quality studies received less than 4 stars. We considered methodological quality when we designed our inclusion criteria table prior to conducting the review, such as the use of validated depression measures, as described in the eligibility criteria table (**Appendix 1**). We resolved all disagreements after discussion.

### Sensitivity analysis

We conducted two sensitivity analyses. First, we conducted sensitivity analyses comparing results from random-effects and fixed-effect models to assess the robustness of pooled estimates according to the Cochrane handbook chapter 10, section 10–14.^53^ Second, we conducted sensitivity analysis that excluded two studies that enrolled “college students” without reporting an explicit age range to assess robustness to potential age misclassification.^54,55^

Given the substantial heterogeneity observed across pooled analyses, we conducted additional sensitivity analyses to assess the robustness of the findings: (1) leave-one-out analyses, (2) geographic subgroup exploration, and (3) prediction interval analyses. For example, for the leave-one-out analyses, we evaluated the influence of individual studies on the pooled estimates. For geographic subgroup exploration, we restricted our meta-analyses to studies conducted in the United States. For the prediction interval analyses, we used the pooled OR or MD, 95% CI, Tau², and df provided in random effect models to estimate the range of true effects that may be expected, and we computed prediction intervals using the metafor package in R Studio (Version 2023.06.1+524).

### Subgroup analysis

We conducted subgroup analyses to explore potential sources of heterogeneity in the association between vitamin D and depression. Subgroups were defined a priori based on biological and demographic factors reported in the literature, specifically sex (male vs. female) and age group (adolescents vs. young adults). Studies were included in subgroup analyses if they provided sex-stratified or age-stratified effect estimates, or sufficient data to calculate them. We interpreted subgroup analyses as exploratory given the limited number of studies available within each subgroup.

### Reporting bias assessment

We used RevMan to create funnel plots for all reported outcomes and performed visual examinations for heterogeneity. However, we were unable to conduct an Egger regression for outcomes reported by less than 10 studies to statistically check for publication bias.^49,56^

### Certainty assessment

The Grading of Recommendations Assessment Development and Evaluation (GRADE) approach was used for certainty assessment for each outcome.^57^ This tool specifies four levels of certainty for a body of evidence for a given outcome: high, moderate, low, and very low. This assessment considers factors such as study design, risk of bias, consistency of results, directness of evidence, and precision of estimates. Two independent blinded reviewers (JN and WC) used GRADE assessment and resolved disagreements.

## RESULTS

### Study selection

Initial searches resulted in 11,309 citations. After removing duplicates and reviewing titles and abstracts, 358 publications remained for full-text review (**Figure 1**). We excluded some citations that did not meet our systematic review inclusion criteria for study design (e.g., commentaries, editorials, or conference abstracts).^58–60^ We included 20 studies in this systematic review.

**Figure 1.**
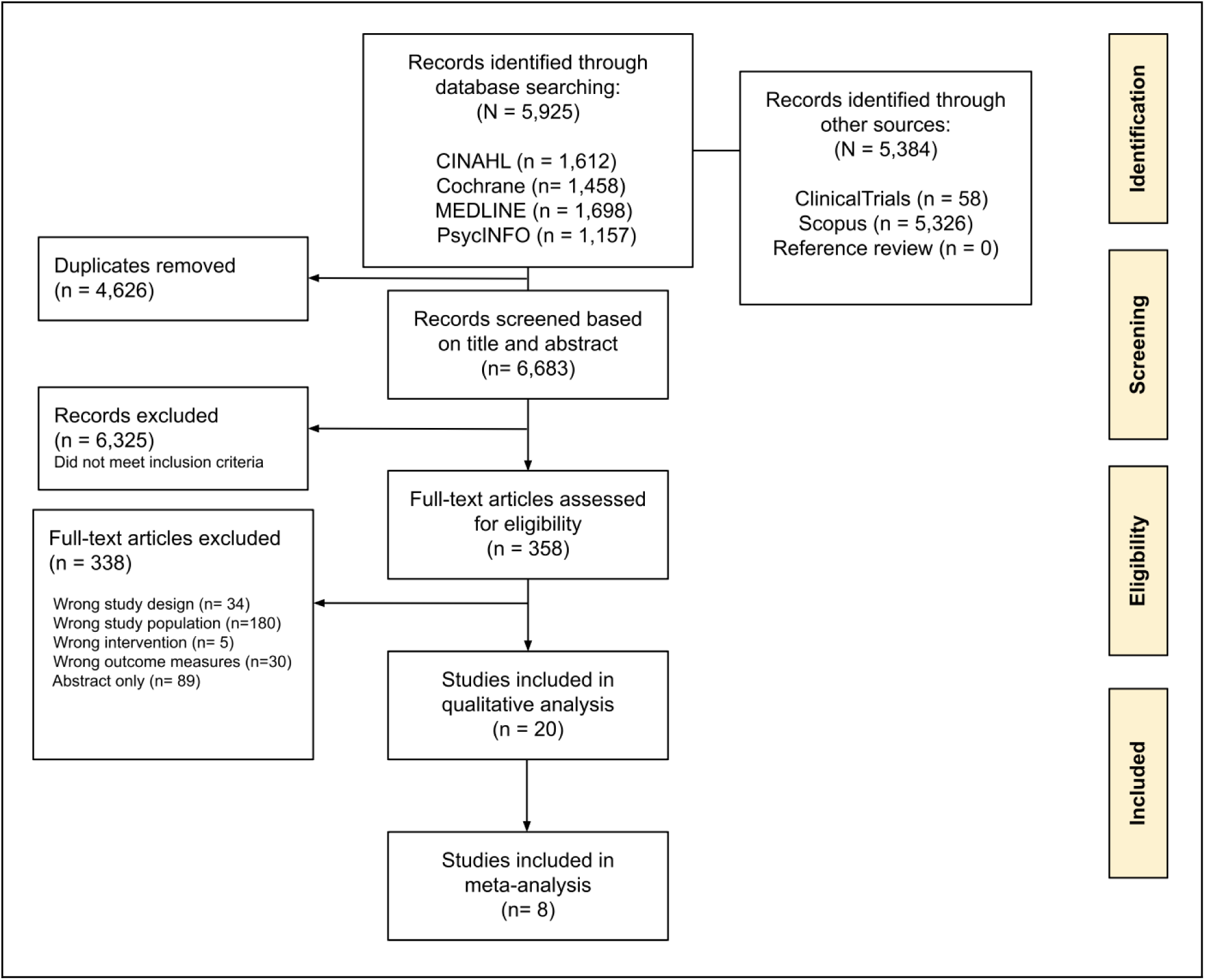
PRISMA flow diagram of search strategy

### Study characteristics

Our review included five cohort studies (25%),^54,61–64^ and 15 cross-sectional studies (75%).^55,65–78^ Included studies were published between 2010 and 2025, with a mean sample size of 27,956 (range: 51 to 483,683 participants) (**Table 1**). Included studies were based in Australia (n=2, 10%), Chile (n=1,5%), India (n=1, 5%), Iraq (n=1, 5%), Iran (n=1, 5%), Korea (n=1, 5%), Kuwait (n=1, 5%), Lithuania (n=1, 5%), New Zealand (n=1, 5%), Saudi Arabia (n=1, 5%), Thailand (n=1, 5%), and the United States of America (n=8, 40%). More than half of the population in the included studies were young adults (18–39 years old) (n=13, 65%), and with variation in ethnicity distribution among their populations. For example, while some studies included participants who were predominantly White,^61,62,68,74–76^ others featured a more diverse sample with a mixed distribution of Black, White, and Asian individuals (**Table 1**).

**Table 1.**
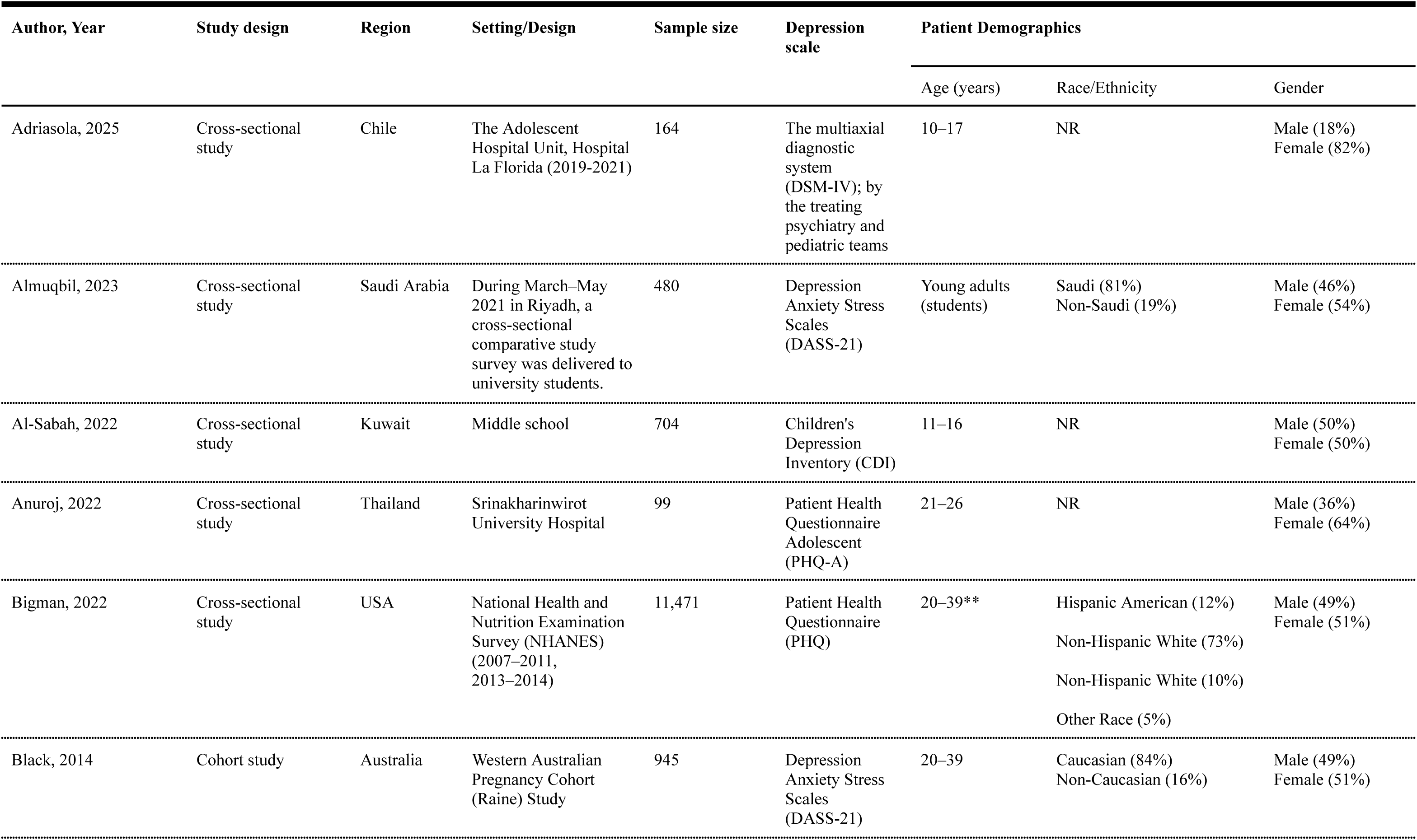
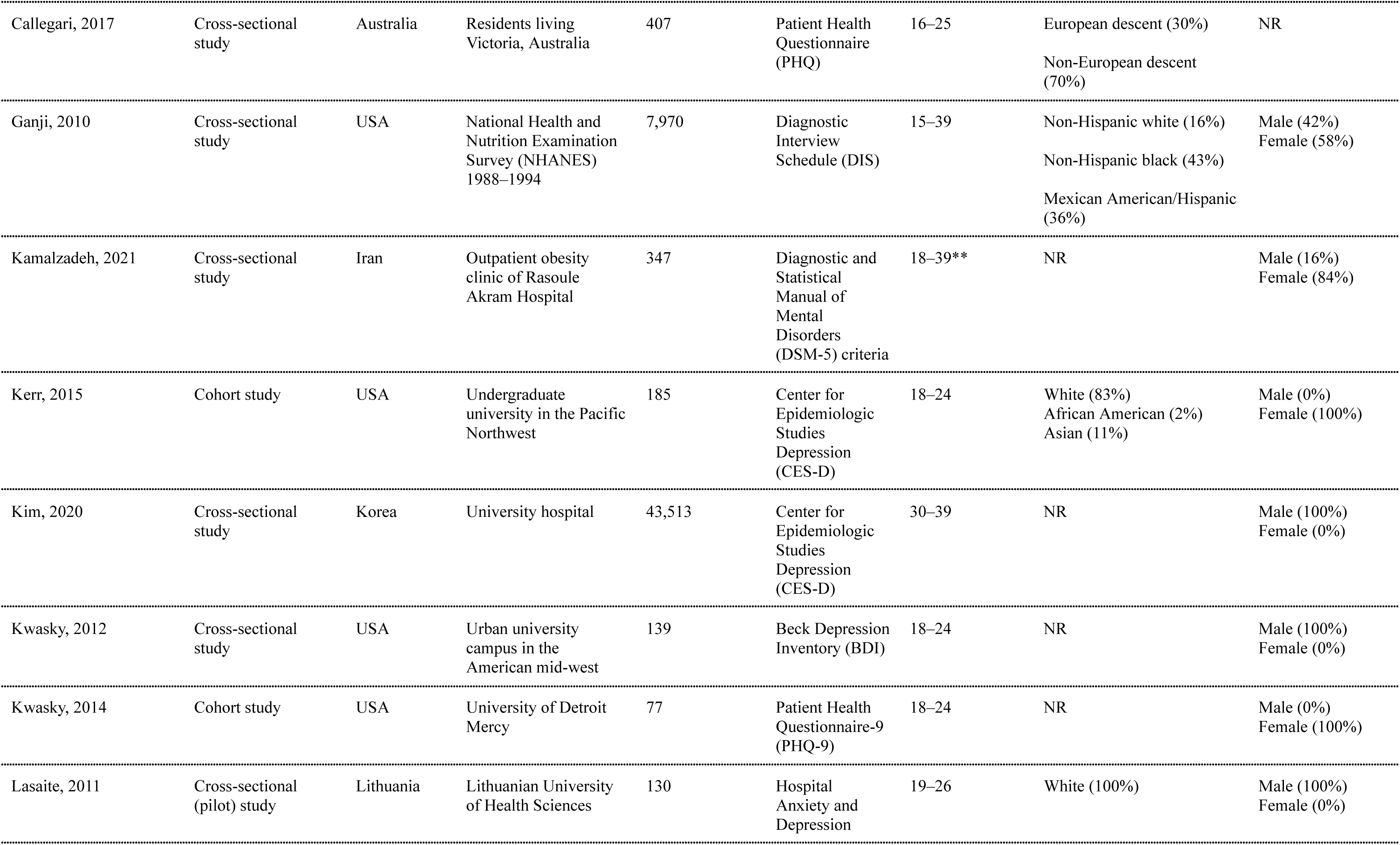
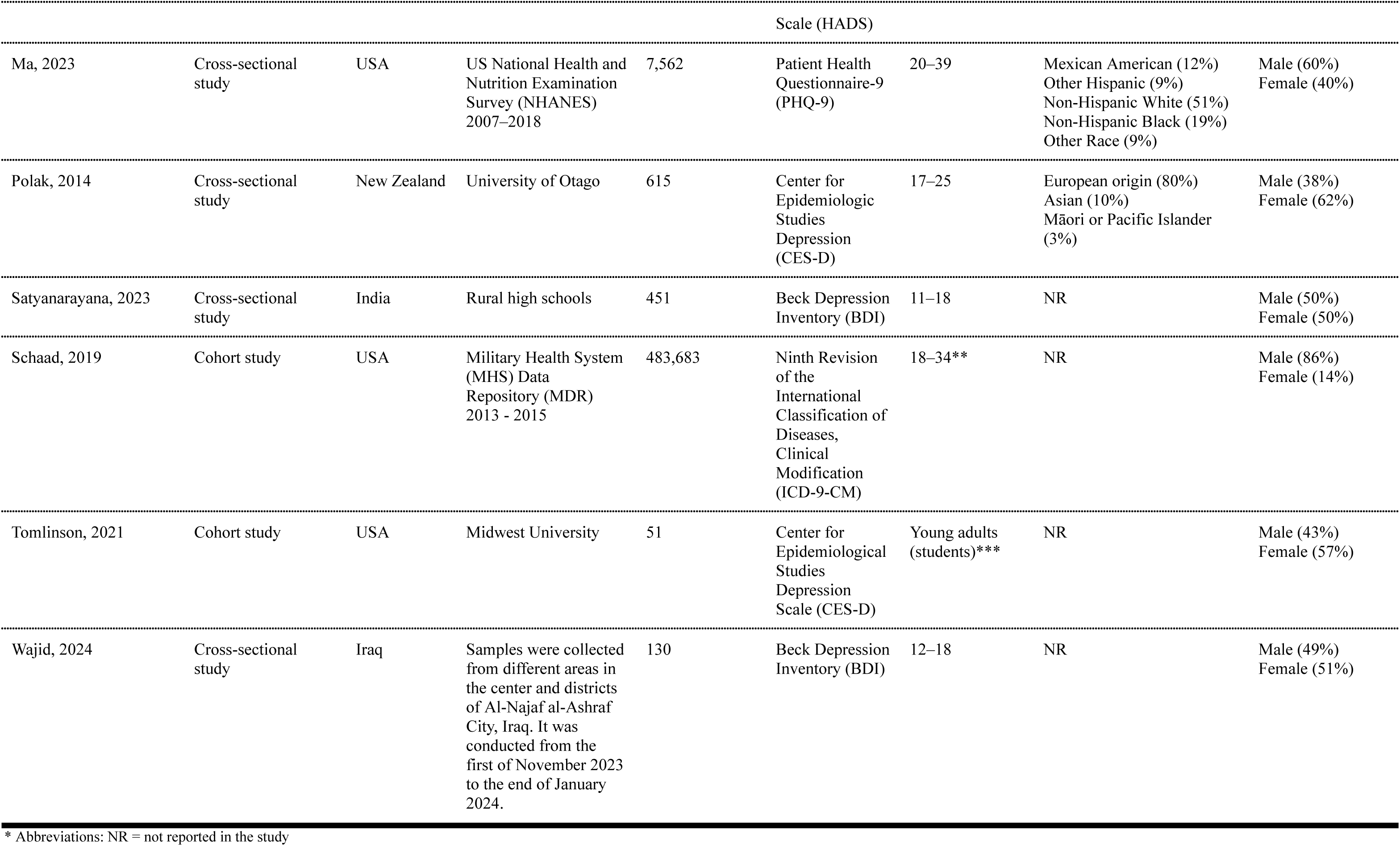
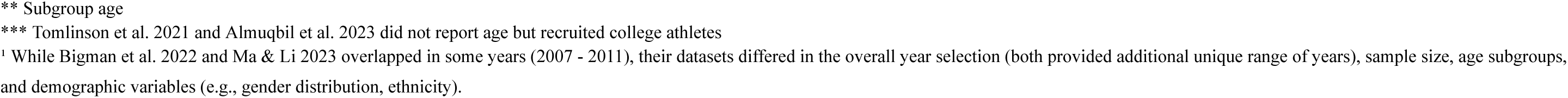
Characteristics of included studies (n=20)

### Risk of bias in studies

All twenty included studies were rated as “good” using the Newcastle-Ottawa Scale to analyze participant selection, comparability, and outcome assessment (**Appendix 6**). Most included studies met the criteria for selection, comparability, and outcome assessment, including the use of validated depression measurement tools and appropriate statistical adjustment for potential confounders. As a result, these studies scored within the range categorized as “good” quality according to the Newcastle–Ottawa Scale criteria.

### Results of individual studies

#### I. Depression measurement instruments

Approximately half of the studies (45%, n=9) used either Patient Health Questionnaire (PHQ) (25%, n=5)^67–69,75^ or Center for Epidemiologic Studies Depression (CES-D) (20%, n=4) (**Table 1**).^54,62,72,76^ Other studies used Children’s Depression Inventory (CDI) (n=1, 5%), Depression Anxiety and Stress Scale (DASS-21) (n=2, 10%), Diagnostic Interview Schedule (DIS) (n=1, 5%), Diagnostic and Statistical Manual of Mental Disorders (DSM-5) (n=1, 5%), Beck Depression Inventory (BDI) (n=4, 20%), the ninth Revision of the International Classification of Diseases, Clinical Modification (ICD-9-CM) (n=1, 5%), or diagnosed by the treating psychiatry team (n=1, 5%) (**Table 1**). All studies tested serum 25(OH)D to measure vitamin D levels and used the United States Institute of Medicine or the Endocrine Society guidelines for the deficiency and insufficiency serum level cutoffs.^39,40,79^

#### II. Association between vitamin D and depression

Of the twenty included studies, twelve studies (60%) after adjustment found that participants with vitamin 25(OH)D_2_ insufficiency (50–75 nmol/L), or deficiency (<50 nmol/L) were more likely to report symptoms of depression (**Table 2**). The remaining eight studies (40%) found no association between vitamin D and depression (**Table 2**). These eight studies had smaller sample sizes (sample size range: 51 to 704) compared to studies that revealed an association (sample size range: 130 to 483,683), and the majority of these studies (n=5 / N=8) were conducted outside of the United States (**Table 1**, **Table 2**). Half (n=4 / N=8) of the studies that reported no association were conducted before 2015. Less than half of cohort studies (n=2 / N=5) found an association between vitamin D and depression.

**Table 2.**
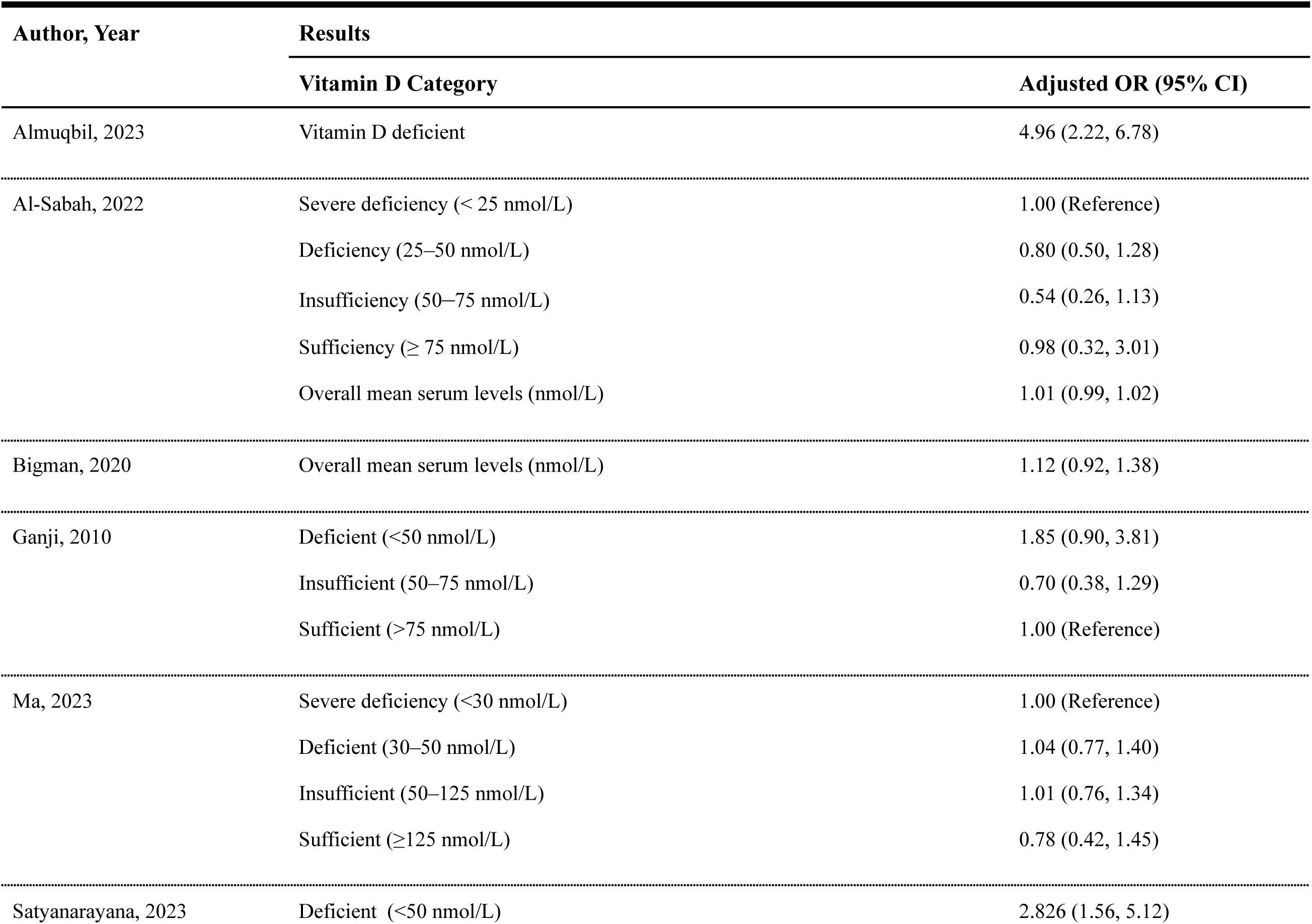
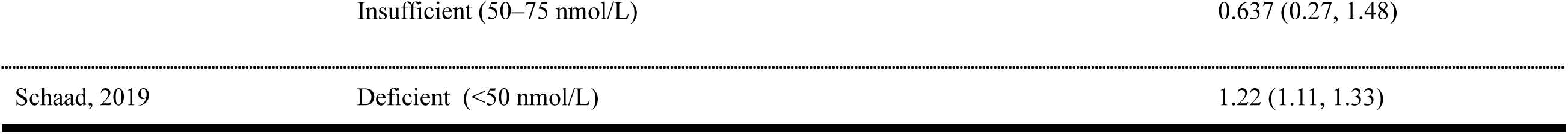
Association between vitamin D serum levels and depression.

Meta-analysis of seven studies with continuous data available revealed that the depressed population of young adults and adolescents had lower mean vitamin D levels than non-depressed participants (Random-effect MD=-17.77 nmol/L [-30.12,-5.42]; p<0.001; I²=95%) (**Figure** 2).^54,70,71,73,75,76^

**Figure 2:**
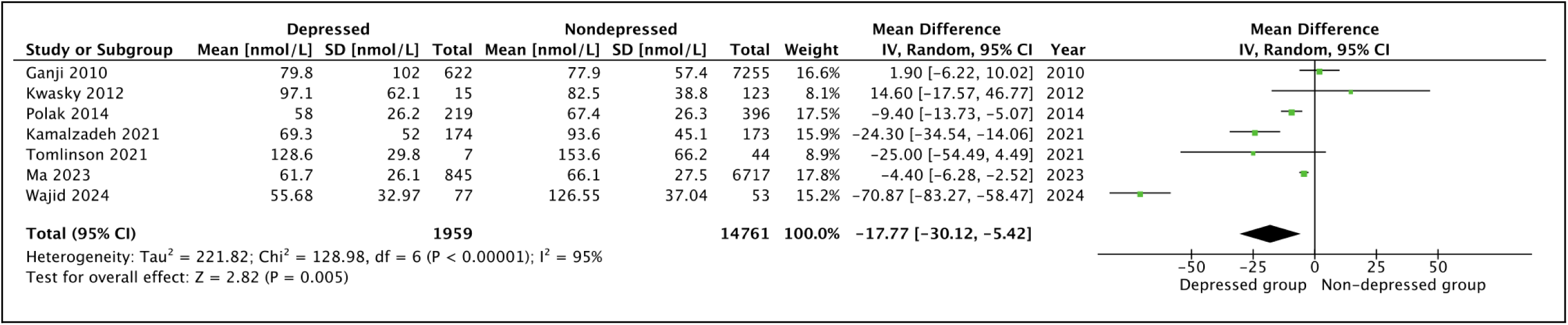
Forest Plot of Association between mean vitamin D serum level (nmol/L) and depression.

For studies that used the vitamin D deficiency level (<50 nmol/L), meta-analysis of five studies that reported odds ratio revealed a higher risk of depression among participants with vitamin D deficiency (Random-effect OR=2.06, 95% CI: 1.02 to 4.17, p<0.001, I^2^=95%) compared to non-depressed participants **(Figure 3A)**.^55,64,66,70,77^

**Figure 3.**
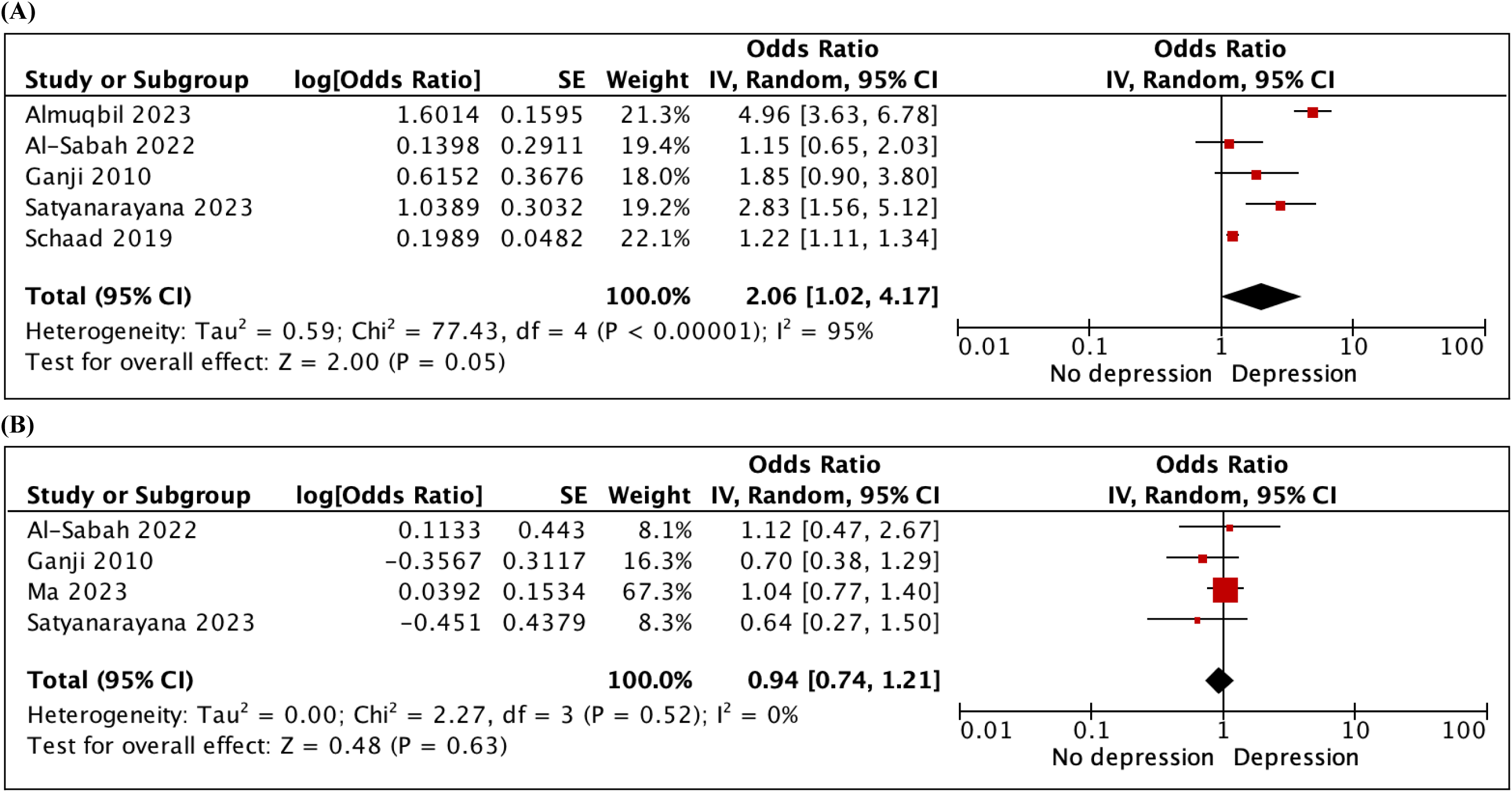
Forest plot of odds ratio (OR) of depression for the lowest v. highest vitamin D categories. Squares to the right of the vertical line indicate that vitamin D deficiency was associated with an increased risk of depression, whereas squares to the left of the vertical line indicate that vitamin D deficiency was associated with a decreased risk of depression. Horizontal lines represent the associated 95% confidence intervals and the diamond represents the overall OR of depression with vitamin D deficiency from the meta-analysis and the corresponding 95% confidence interval. **(A)** vitamin D deficiency and depression and **(B)** vitamin D insufficiency and depression.

On the other hand, for studies that used the vitamin D insufficiency level (50–75 nmol/L), meta-analysis of four studies that reported odds ratio on the association between vitamin D insufficiency levels and depression revealed no significant increase or decrease in risk of experiencing depression (Random-effect, OR = 0.94, 95% CI: 0.74 to 1.21, p = 0.52, I^2^ = 0%) (Figure 3B).^66,70,75,77^

### Sensitivity analysis

#### I. Model comparison

Our sensitivity analysis revealed consistent results across meta-analyses when we compared fixed-effect vs. random-effect models and when we excluded studies that report college students’ age in their studies **(Appendix 8A–8I)**.

#### II. Leave-one-out sensitivity analyses

Our leave-one-out sensitivity analyses for all reported meta-analyses revealed that removal of any single study did not alter the overall effect estimates and statistical significance of the pooled results. Also, our exclusion of cohort studies from meta-analyses that included both cross-sectional and cohort studies did not significantly change our overall findings. One study used non-standard vitamin D category cutoffs for insufficiency that could not be fully reclassified according to our predefined thresholds. However, leave-one-out sensitivity analyses indicated that its exclusion did not have a significant impact the pooled results.^75^

#### III. Geographic subgroup exploration

When we restricted the meta-analysis of studies reporting mean differences in vitamin D serum levels (**Figure 2**) to four studies conducted in the United States, we found that heterogeneity was significantly reduced (I² = 45%) compared with the overall analysis (I² = 95%) (**Appendix 8J)**.^54,70,73,75^ In this subgroup analysis, we found that the pooled mean difference between depressed and nondepressed participants was no longer statistically significant −2.57 nmol/L (95% CI: −9.31 to 4.17) (**Appendix 8J**).

#### III. Prediction Interval

Given the high heterogeneity observed across pooled analyses, we computed 95% prediction intervals. For **Figure 2**, the pooled mean difference in vitamin D serum levels had a 95% prediction interval of −59.34 to 23.8. For the odds ratio meta-analyses, we found between-study variability with prediction intervals that ranged from 0.14 to 30.61 for **Figure 3A**, 0.46 to 7.72 for **Figure 4A**, and 0.01 to 104.67 for **Figure 4B**.

**Figure 4.**
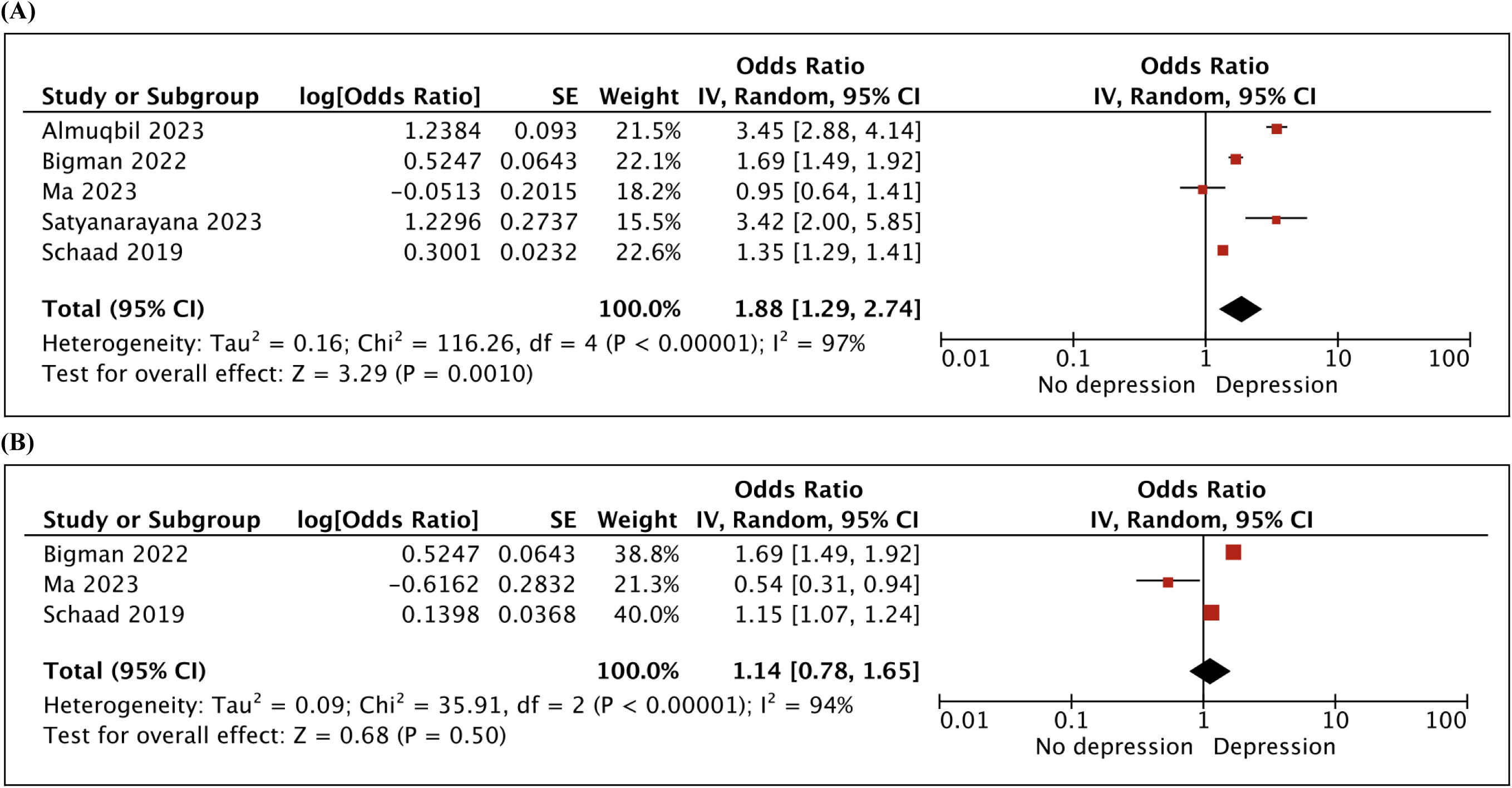
Forest plot of odds ratio of association between vitamin D and depression among **(A)** female participants compared to male participants and **(B)** young adult (age range: 25–39 years old) participants compared to other age group participants.

### Subgroup analysis

#### I. Sex differences in t he association between vitamin D and depression

Seven studies investigated sex differences in the association between vitamin D and depression (**Table 3**).^55,61,64,68,71,75,77^ One study reported the results in rate ratio and found sex differences in the association between vitamin D and depression.^61^ This study found a significant association between vitamin D serum concentration and symptoms of depression in males but no significant associations in females. Another study reported results using computed mean and standard deviation values and found that while the mean serum vitamin D levels was lower in depressed male participants compared to depressed female participants, the difference was not statistically significant.^71^

**Table 3.** Age and gender disparities in association between vitamin D and depression for meta-analysis.

| Author, Year | Results |  |
| --- | --- | --- |
|  | Category | Adjusted OR (95% CI) |
| **Gender** |  |  |
| Almuqbil, 2023 | *Male* | 1.00 (Reference) |
|  | *Female* | 3.45 (1.99, 4.14) |
| Bigman, 2022 | *Male* | 1.00 (Reference) |
|  | *Female* | 1.69 (1.49, 1.91) |
| Ma, 2023 | *Male* | 0.65 (0.42, 1.01) |
|  | *Female* | 0.57 (0.40, 0.82) |
| Satyanarayana, 2023 | *Male* | 1.00 (Reference) |
|  | *Female* | 3.42 (2.00, 5.85) |
| Schaad, 2019 | *Male* | 1.00 (Reference) |
|  | *Female* | 1.35 (1.29, 1.40) |
| **Age** |  |  |
| Bigman, 2022 | *20–29 years* | 1.00 (Reference) |
|  | *30–39 years* | 1.12 (0.92, 1.38) |
| Ma, 2023 | *29–39 years* | 0.54 (0.31, 0.95) |
| Schaad, 2019 | *18–24 years* | 1.13 (1.04, 1.24) |
|  | *25–34 years* | 1.15 (1.07, 1.25) |
|  | *35–44 years* | 1.14 (1.05, 1.23) |

We conducted a meta-analysis on five studies that reported an odds ratio on the association between vitamin D and depression and found an increased risk of experiencing depression among female participants compared to male participants (Random-effect, OR=1.88, 95% CI: 1.29 to 2.74, p<0.001; I^2^=97%) (**Figure 4A).**^55,64,68,75,77^ More than half of these studies (n=3, 60%) reported equal distribution of male and female participants (**Table 1**) (**Table 2**).^55,68,77^

#### II. Age differences in the association between vitamin D and depression

Three studies investigated age differences in the association between vitamin D and depression,^64,68,72^ and two studies reported an increase in the odds of having a strong association as participants age (**Table 3**). Meta-analysis of three studies that reported odds ratio on the association between vitamin D levels and depression among young adults (age range: 25-39 years old) was not significant and uniform across all three studies (Random-effect OR = 1.14, 95% CI: 0.78 to 1.65, p < 0.001, I^2^ = 94%) (**Figure 4B).**^64,68,75^

### Certainty assessment

We created a GRADE certainty rating for 5 meta-analysis outcomes: (1) mean difference of vitamin D serum levels (depressed vs. non-depressed), (2) OR for vitamin D deficiency, (3) OR for vitamin D insufficiency, (4) OR for vitamin D female subgroup, and (5) OR for vitamin D young adults aged 25–39 years old. We found certainty assessment to be “very low” across all 5 outcomes.

## DISCUSSION

### Key findings

The findings of our systematic review and meta-analysis indicate that vitamin D deficiency is associated with depression among adolescents and young adults. There is substantial heterogeneity across studies. Three significant results were revealed in this review: (1) participants who had deficiency (<50 nmol/L) in vitamin D serum levels were likely to have more severe depression, (2) female participants were more likely to have an association between vitamin D deficiency and depression compared to male participants, and (3) there is no association between vitamin D and depression among young adults aged 25–39 years.

### Results in context

Overall, the findings of our review agree with the evidence from previous studies and fill gaps in knowledge regarding how vitamin D deficiency contributes to depression. Our review included several published studies that were not included in two previous systematic reviews on this topic,^80,81^ and all but one recently published study found a significant association between vitamin D deficiency and depression.^54,62,64,68,71,72,75,77^ Other population-based studies focusing on later stages of adulthood have demonstrated a significant association between vitamin D deficiency and depression.^30,80^ While more than half of our studies found a significant association between vitamin D deficiency and depression, the rest of the studies were consistent with previously published studies that found no association between vitamin D deficiency and depression.^24–27^ Therefore, our results do not necessarily resolve the controversy, but provide insight into the association of vitamin D deficiency and depression, specifically among adolescents and young adults. Furthermore, compared to individuals who have vitamin D deficiency (<50 nmol/L), we found that individuals with vitamin D insufficient serum levels (50–75 nmol/L) may not experience a lower risk of depressive symptoms. This may be explained through several biological pathways, as vitamin D has been shown to influence serotonin synthesis, regulate neuroinflammatory processes, and interact with vitamin D receptors in brain regions that play a critical role in mood regulation.^82–84^ Our results regarding the contribution of sex and age disparities in this association between vitamin D deficiency and depression are consistent with previous studies that focused on sex and age disparities in depression.^85,86^ Furthermore, these observed sex differences may be explained by biological, psychosocial, and behavioral differences that may affect mental health.^87,88^

### Quality of the evidence

All included studies used the United States Institute of Medicine Vitamin D guidelines for the deficiency and insufficiency serum level cutoff. Also, studies used various validated measures for depression. However, some studies had a small sample size (e.g., n < 100) relative to other studies included, which can limit the quality of evidence by increasing the risk of Type I errors (false positives) and Type II errors, and limiting the generalizability of the findings. Furthermore, a larger sample size generally enhances the reliability and validity of the study’s findings and increases the chance of detecting an association between vitamin D deficiency and depression. In addition, the smaller number of studies (n = 8) available for meta-analysis may limit the overall strength and precision of the pooled estimates, as more extensive meta-analyses generally provide more reliable conclusions. Although random-effects models were used to account for between-study variability, the high I² values and wide prediction intervals suggest that the magnitude of the association between vitamin D serum levels and depression may vary across different study regions. Therefore, the overall findings were exploratory and should be interpreted with caution.

### Strengths and limitations

Our systematic review consists of recently published research studies investigating the association between vitamin D deficiency and depression among adolescents and young adults. We found studies conducted in the United States and overseas, which increases generalizability of the results. Another key strength of this systematic review is the robust assessment of methodological quality, with all included studies rated as ‘good’ based on the Newcastle-Ottawa Scale, ensuring high reliability and rigor in the findings. However, one limitation is that studies conducted outside of the United States may introduce cultural biases, which could affect the perception and reporting of mental health issues, potentially leading to variations in the results and limiting their generalizability to the U.S. population. Another limitation is that our systematic review consists of studies with a sample size ranging from small to large-scale populations, which may affect the consistency of the findings. Also, our study does not indicate the direction of the effect between vitamin D deficiency and depression. Since most included studies were cross-sectional, we cannot establish temporality and rule out reverse causation. For example, our systematic review and meta analysis does not reveal whether low vitamin D serum levels cause depression or whether depression causes low vitamin D serum levels. Furthermore, because relatively few studies met inclusion criteria for meta-analysis among this population of interest (n= <10), we were unable to perform formal statistical tests for publication bias, and the possibility of selective publication cannot be excluded.

### Impact, implications, and next steps

The results of this systematic review indicated that there is evidence suggesting vitamin D deficiency is associated with an increase in an individual’s likelihood of having depression. Based on the findings, the results may impact adolescents (aged 10–17 years) and young adults (aged 18–39 years), parents and families of these individuals, schools, healthcare providers, pharmaceutical companies, public policymakers, and researchers. The implications for this review may include the potential to reevaluate how depression is currently screened, as there may be an opportunity to incorporate an examination of current vitamin D levels more significantly among this population. This may thereby impact healthcare professionals and potentially restructure standard paths of care in this area. Additionally, the associations found within this target population having depression could further impact potential risk factors that healthcare professionals should be mindful of when conducting screenings, such as the patient’s gender.

Our findings emphasize the need for greater attention to vitamin D status among younger populations, who are often overlooked in depression research. Although current evidence does not permit definitive clinical recommendations, identifying and correcting vitamin D deficiency remains a reasonable preventive measure given its established role in bone and immune health. When we restricted our sensitivity analyses to studies conducted in the United States, we found a significantly reduced heterogeneity, which suggests that geographic differences in population characteristics may partially explain the variability observed in our meta-analyses. Also, our leave-one-out sensitivity analyses suggest that our overall meta-analysis findings were not driven by any individual study. Furthermore, our systematic review revealed that the association between vitamin D and depression may not be universally generalizable across populations and could be influenced by country-specific and environmental factors that affect serum vitamin D levels. For example, vitamin D serum levels could be influenced by sunlight exposure during outdoor activities, which may vary across countries and populations.

Depression is a complex condition influenced by biological, psychological, social, and environmental factors (**Figure 5**). While our review focused on *v*itamin D, which represents a critical biological factor within the nutritional and physiological determinants of mental health, it likely plays a complementary role alongside other nutrients and environmental influences that contribute to the development of depression.^89^

**Figure 5.**
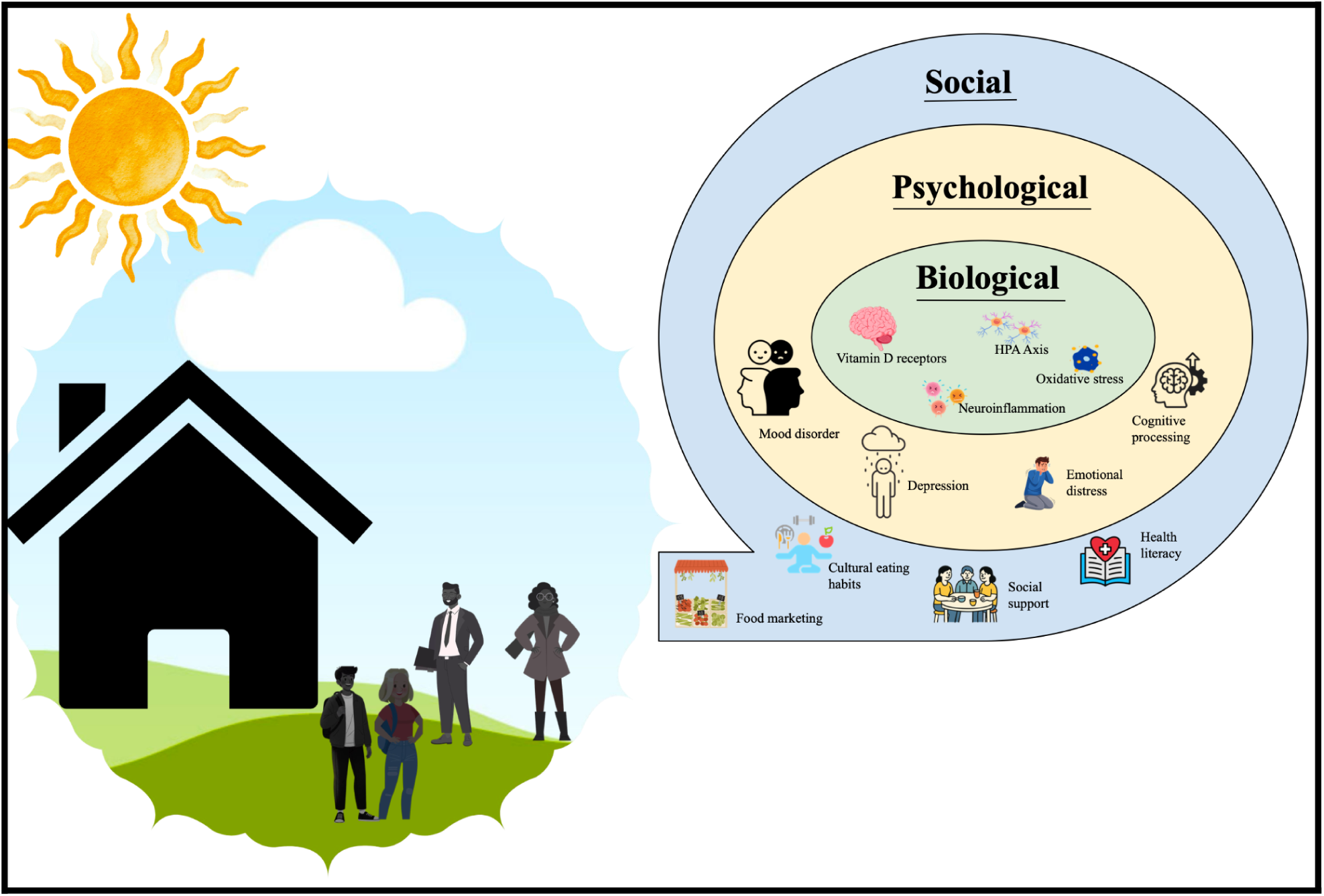
Conceptual framework illustrating the multifactorial determinants of depression and the potential role of vitamin D among adolescents and young adults

Given the low certainty of evidence based on the GRADE assessment, our findings should be considered hypothesis-generating. Additional well-designed prospective studies and randomized controlled trials will be helpful to investigate the causal relationship between vitamin D status and depression and whether optimizing vitamin D levels can reduce depression risk or improve mood outcomes. In addition, public health strategies that promote safe sun exposure, dietary fortification, and awareness of vitamin D deficiency may have downstream mental health benefits.

Future research based on the results of this systematic review may include randomized clinical trials to examine if similar strong associations are demonstrated with vitamin D supplementation. Furthermore, follow-up research may further explore the impact of age and sex on the association between vitamin D deficiency and depression using a larger number of studies and a more diverse populations (e.g., race and ethnicity), as adolescence and young adulthood represent a critical period for mental health and there are well documented gender disparities in the prevalence and presentation of depression among this group.^85,90^ Also, future studies may conduct subgroup analyses based on latitude, UV index, or season of blood sampling to investigate whether environmental factors influence the association between vitamin D serum levels and depression.

## CONCLUSION

In summary, our meta-analyses revealed an association between vitamin D deficiency and depression among adolescents and young adults. Our results suggest a possible association, with exploratory analyses indicating potential differences by sex and age in the association between vitamin D and depression. Given the high heterogeneity across studies, these findings should be considered cautiously. Follow-up prospective studies and randomized controlled trials with vitamin D supplementation are critical to further explore the temporal and causal association between vitamin D status and depression.

## Review Protocol

We created a protocol in February 2023 to guide the inclusion of studies. We registered our protocol in November 2023 (https://osf.io/ujrt4) (See **Appendix 7** for systematic review protocol modifications).

## Data Availability Statement

The data used and analyzed in this systematic review are available upon request from the corresponding author.

## CRediT authorship contribution statement

**Joseph P. Nano:** Conceptualization, methodology, formal analysis, writing – original draft. **William Catterall:** methodology, formal analysis, writing – original draft**. Violet Filer:** Conceptualization, methodology, writing – review and editing**. Salar Khaleghzadegan:** methodology, writing – review and editing. **Ricardo Almazan Jr.**: Conceptualization, writing – review and editing. **Heather B. Blunt:** methodology, writing – review and editing. **Renata W. Yen:** Conceptualization, methodology, formal analysis, writing – original draft.

## Acknowledgments

We would like to thank the following faculty and colleagues of The Dartmouth Institute for Health Policy and Clinical Practice for their feedback and contributions to the development of this research topic: Dr. Amber Barnato, Dr. Honor Passow, E. Chandlee Bryan, Elaina Vitale, Elizabeth A. Koelsch, Roland Lamb, and Vrushabh Ladage.

## APPENDICES

**Appendix 1:** Inclusion criteria table & outcomes of interest.

| Category | Inclusion Criteria | Justification |
| --- | --- | --- |
| Study design | Cohort study | Cohort study can investigate the association between multiple outcomes and exposures in a single study for a long period of time. This is important to show changes in vitamin D serum concentration and depression scores over time. |
|  | Case study | Case studies focus on a single individual or group and allow researchers to collect information on unusual cases that could be related to association between vitamin D and depression. |
|  | Cross-sectional | Cross-sectional studies provide comparison between different variables, such as sleep, age, LDL levels, vitamin D serum concentration, and depression scores at the same time. Survey study designs are observational as there is no experimental component. Also, they are useful in recruiting a large number of participants and describing the characteristics of a large population, such as age, race, hours of sleep, and past medical history. |
|  | Case-control studies | Case-control studies compare vitamin D levels between individuals with and without depression, allowing estimation of the association between exposure and outcome. This design is particularly valuable when investigating conditions with relatively low prevalence (e.g., diagnosed depression in younger populations). |
|  | Pilot study | Pilot study can be used to provide information about association between vitamin D and depression by conducting a smaller scale study to assess the safety of treatment or interventions.<sup>91</sup> |
|  | Randomized control trial study | Randomized control trials (RCT) can be used to investigate whether there is a causal relationship between vitamin D and Depression, and reduce chances of selection bias.<sup>92</sup> |
| Population | Adolescents <sup>33,80,93</sup> (defined by $\geq 10$ and $17 \leq$) and young adults <sup>94,95</sup> ($\geq 18$ and $\leq 39$) | This is the most relevant population to our research topic, because depression is more likely to start during adolescence and this population is less likely to experience morbidity that would increase the risk for depression.<sup>96</sup> If feasible, we would plan to evaluate studies of young adults ($\geq 18$ and $\leq 39$), and perhaps a separately subgroup-analysis of adolescents ($\geq 10$ and $17 \leq$) to extend our findings. The age cutoff for adolescents was based on the World Health Organization's age standard.<sup>33</sup> |
| Intervention / Exposure | Serum Vitamin D deficient ($< 50$ nmol/L) and insufficient ($50-75$ nmol/L) | This is based on the United States Institute of Medicine Vitamin D cutoff.<sup>22,40,79,97</sup> |
| Comparison condition | Serum Vitamin D sufficiency ($> 75$ nmol/L) | This is based on the United States Institute of Medicine Vitamin D cutoff.<sup>22,40,79,97</sup> |
| Outcome | Depression | Several depression questionnaires are used to measure depression (**Appendix 5**). |

**Appendix 2:** PubMed Search Terms Table.

| Theme | MeSH terms (exploded) | Key words |
| --- | --- | --- |
| Depression | "Depression"[Mesh] OR<br>"Depressive Disorder"[Mesh] | Depression [TIAB] OR Depressive Disorder [TIAB] OR<br>seasonal affective[TIAB] OR Dysthym*[TIAB] OR<br>affective disorder*[TIAB] OR major depress*[TIAB] OR<br>unipolar depress*[TIAB] |
| Young Adults and<br>Adolescent | "Young Adult"[Mesh] OR<br>"Adolescent"[Mesh] | Young Adult* [TIAB] OR Adolescen* [TIAB] OR Teen*<br>[TIAB] |
| Vitamin D | "Vitamin D"[Mesh] OR "Vitamin D<br>Deficiency"[Mesh] OR<br>Hydroxycholecalciferols[Mesh] OR<br>cholecalciferol[Mesh] OR<br>ergocalciferol[Mesh] | Vitamin D* [TIAB] OR hydroxycholecalciferol*[TIAB]<br>OR 25-hydroxyvitamin D[TIAB] OR<br>cholecalciferol*[TIAB] OR ergocalciferol*[TIAB] OR<br>calcifediol*[TIAB] OR calcitriol*[TIAB] OR<br>hydroxyvitamin*[TIAB] |

**Appendix 3:**
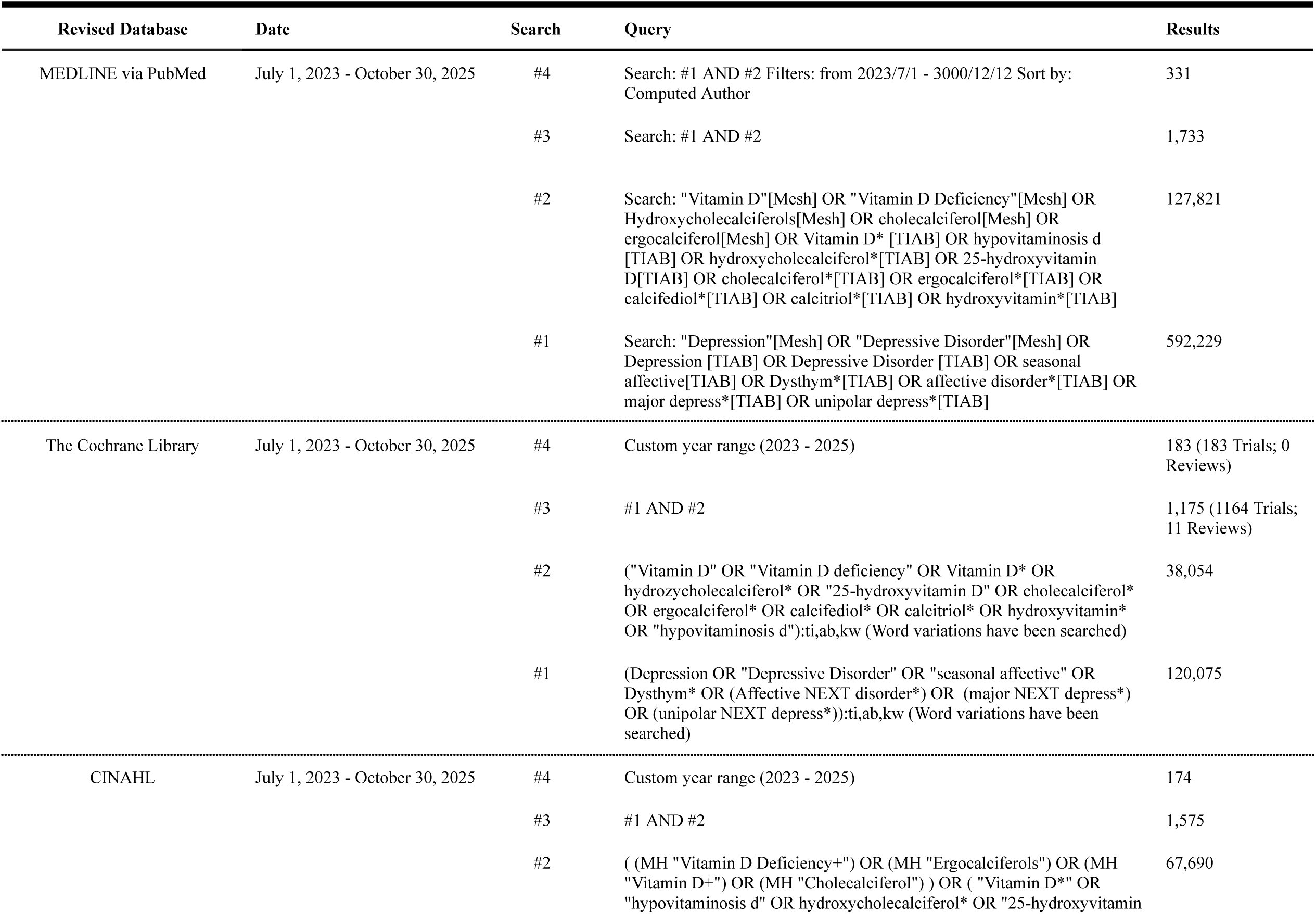
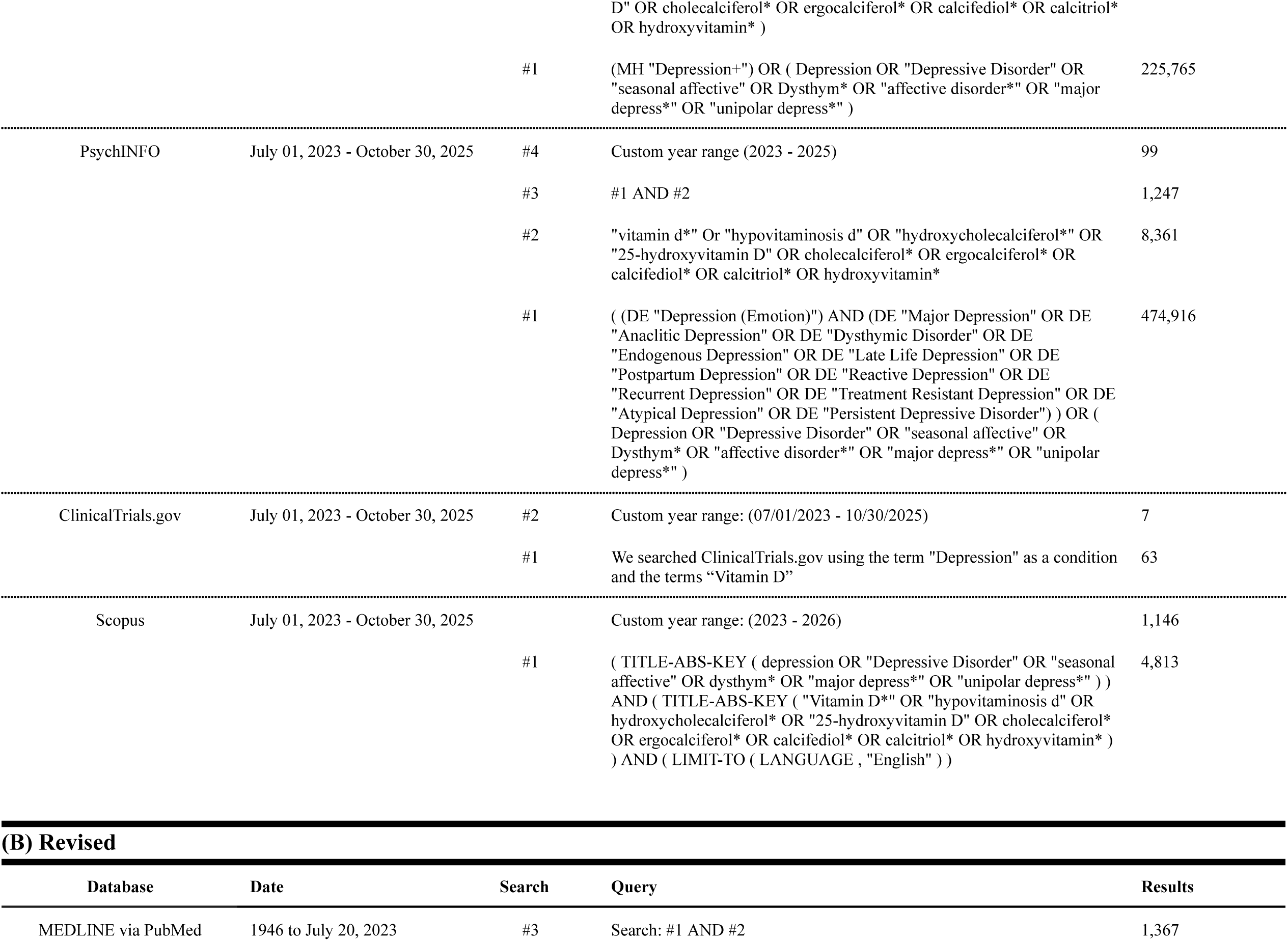
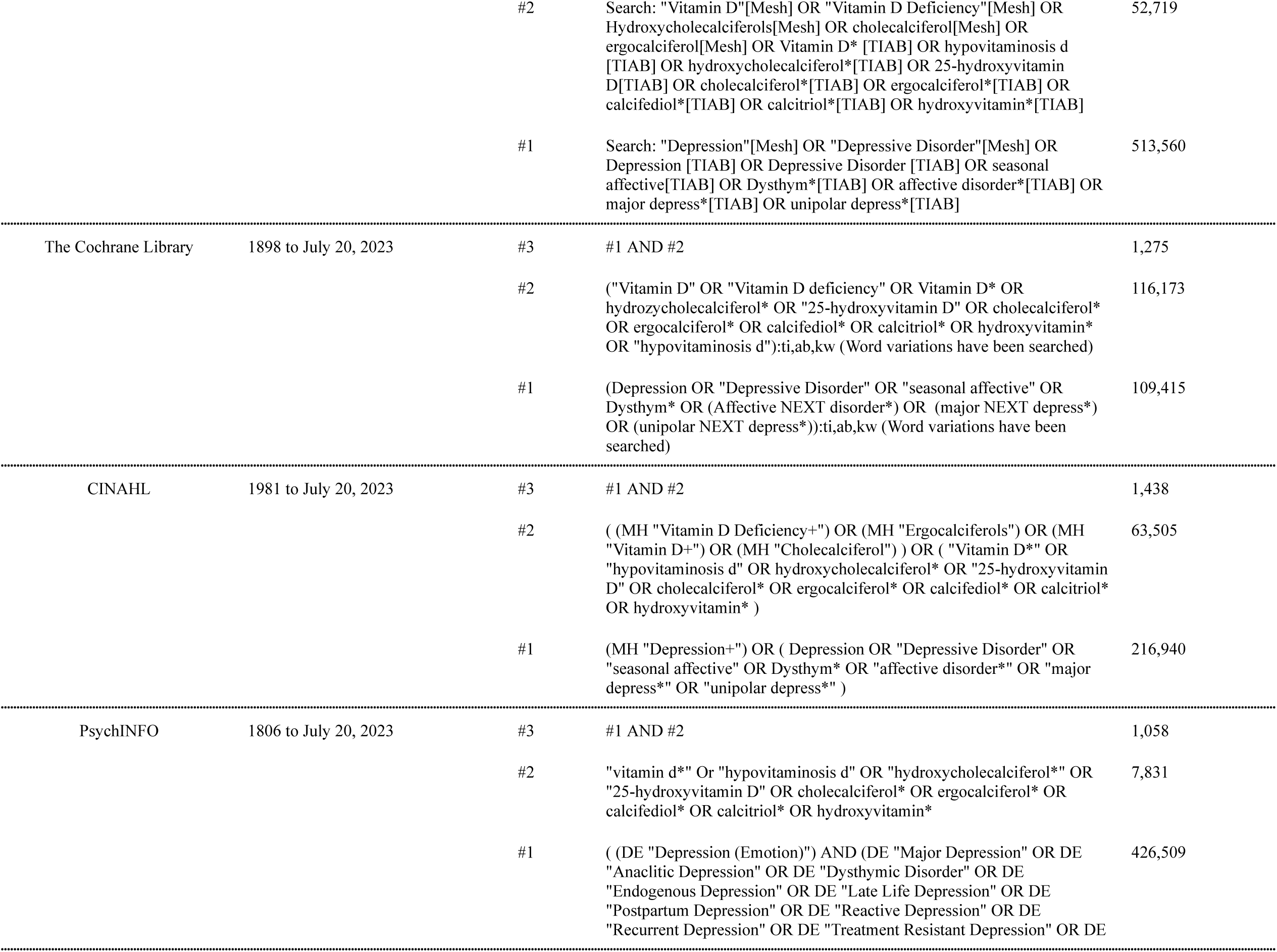
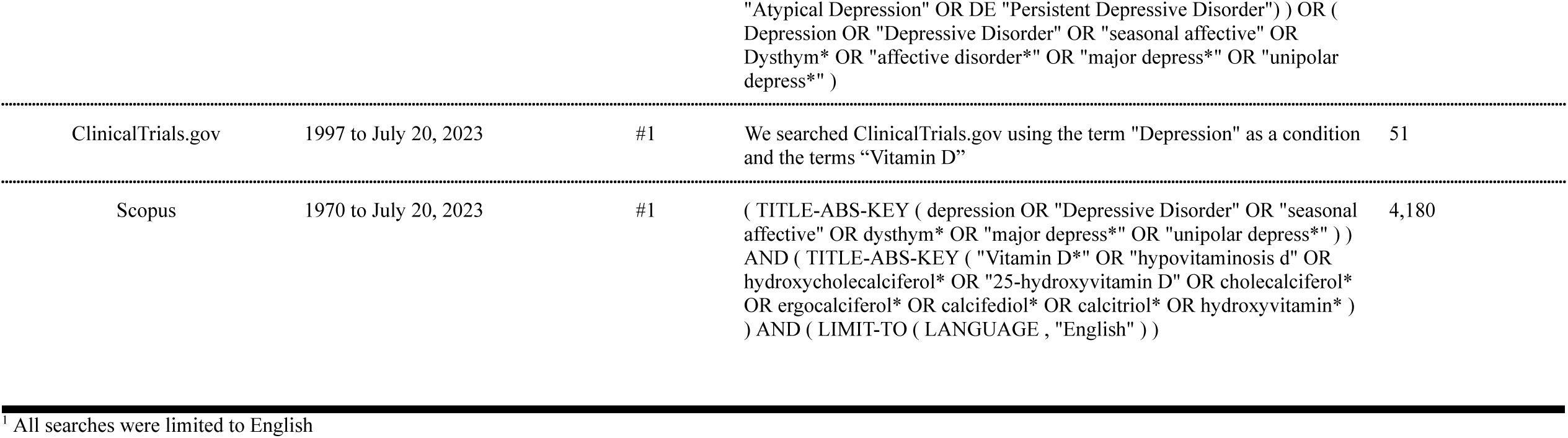
Search Strategies (A) Updated and (B) Original^1^.

**Appendix 4:** Data Collection Form.

| First author | Publication year | Setting / Database | Study design | Sample size | Demographics | Primary outcome | Secondary outcome | Key conclusion | Extractor comment |
| --- | --- | --- | --- | --- | --- | --- | --- | --- | --- |
| ..... |  |  |  |  |  |  |  |  |  |

**Appendix 5.**
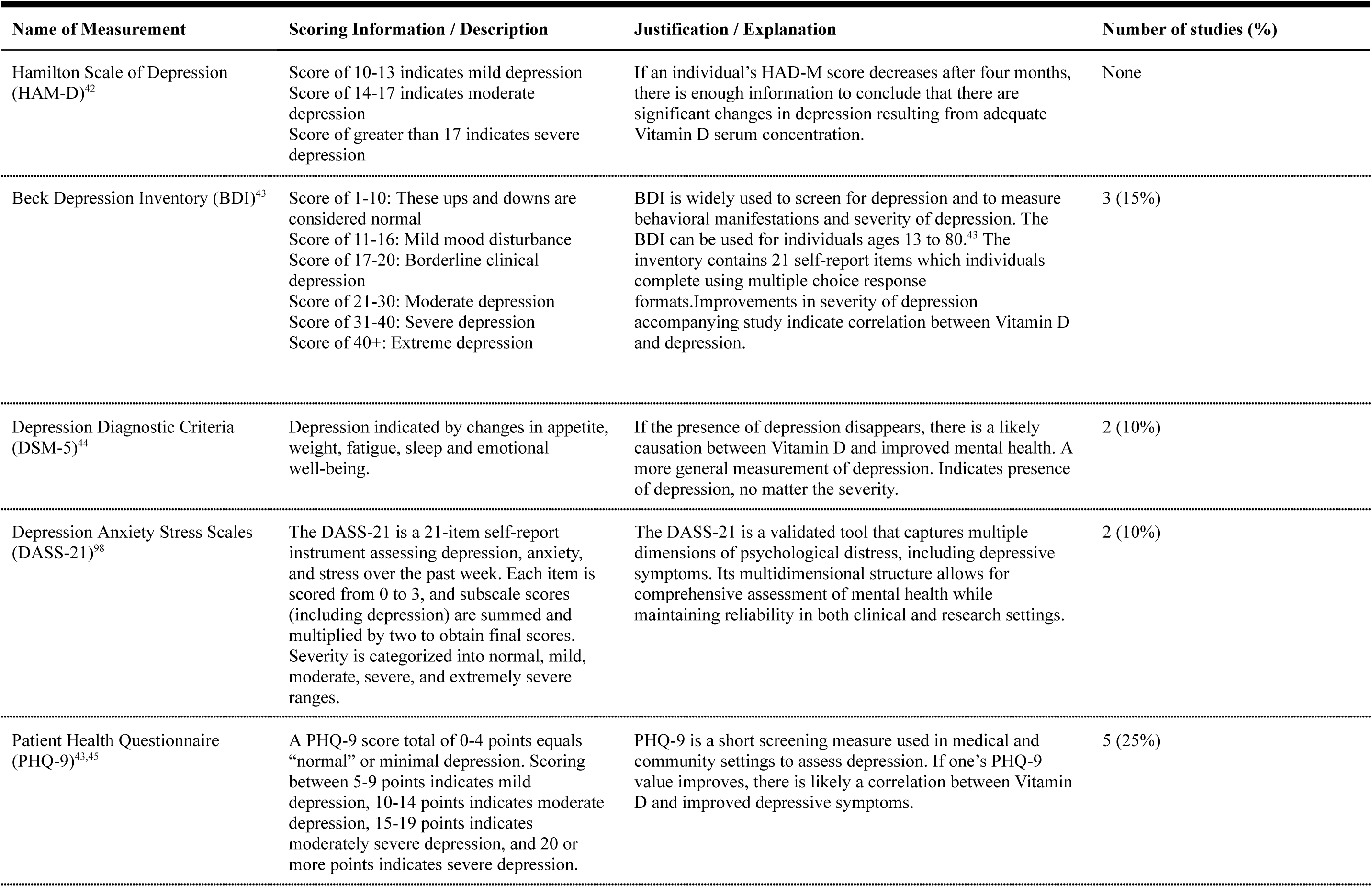
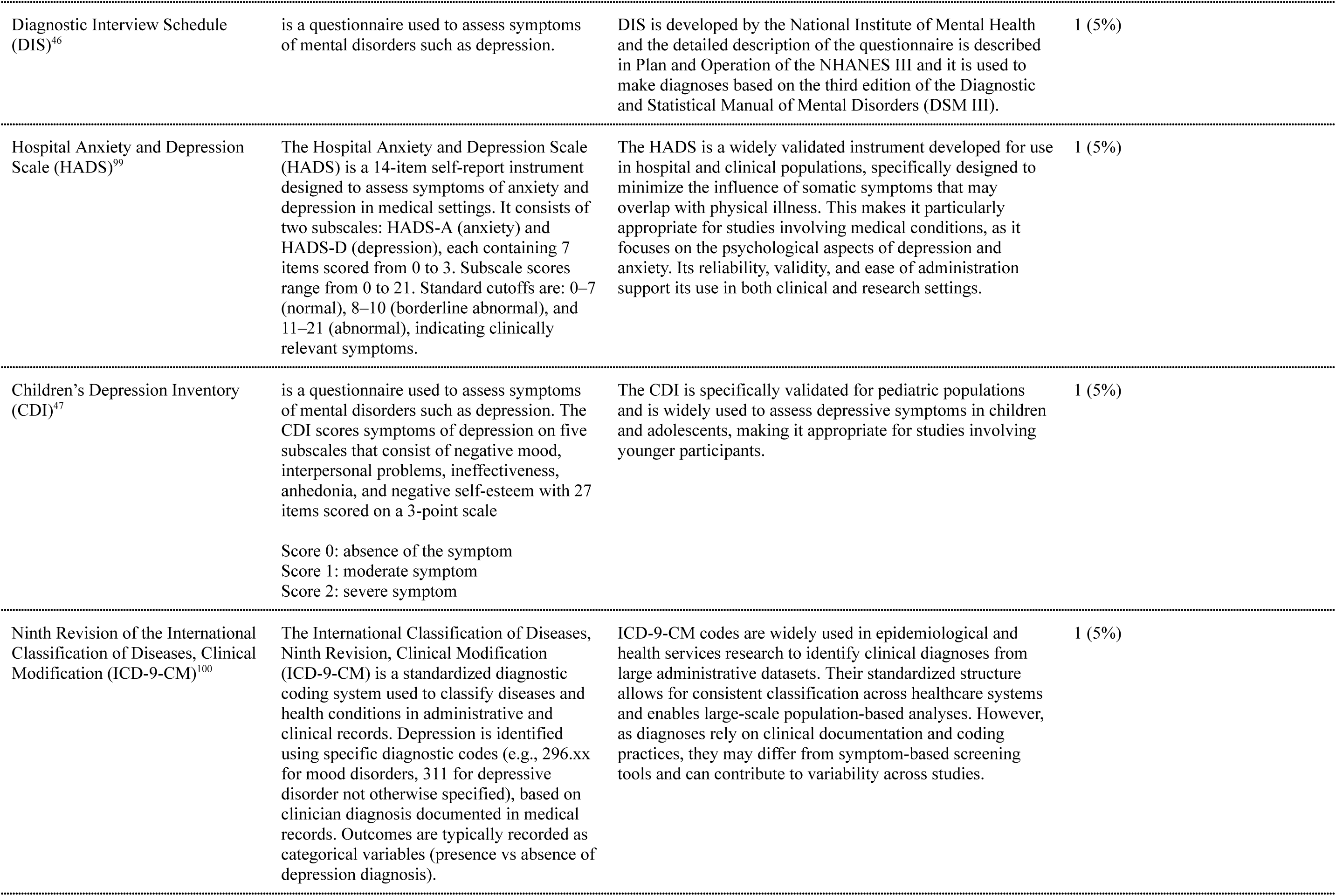
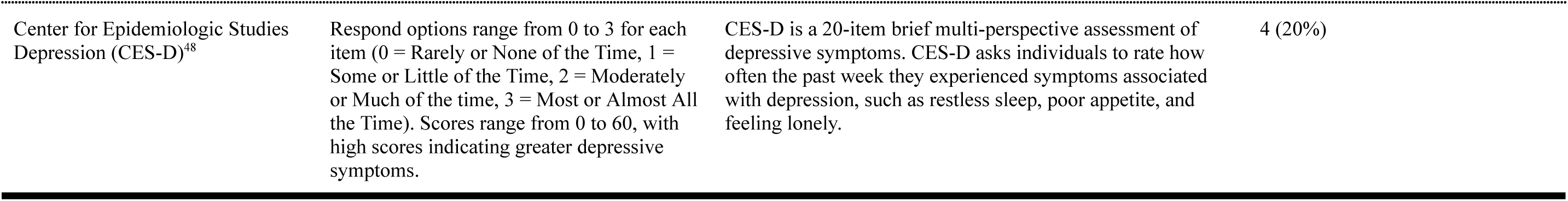
Summary of primary outcome measurements.

| Name of Measurement | Scoring Information / Description | Justification / Explanation | Number of studies (%) |
| --- | --- | --- | --- |
| Hamilton Scale of Depression (HAM-D)<sup>42</sup> | Score of 10-13 indicates mild depression<br>Score of 14-17 indicates moderate depression<br>Score of greater than 17 indicates severe depression | If an individual's HAD-M score decreases after four months, there is enough information to conclude that there are significant changes in depression resulting from adequate Vitamin D serum concentration. | None |
| Beck Depression Inventory (BDI)<sup>43</sup> | Score of 1-10: These ups and downs are considered normal<br>Score of 11-16: Mild mood disturbance<br>Score of 17-20: Borderline clinical depression<br>Score of 21-30: Moderate depression<br>Score of 31-40: Severe depression<br>Score of 40+: Extreme depression | BDI is widely used to screen for depression and to measure behavioral manifestations and severity of depression. The BDI can be used for individuals ages 13 to 80.<sup>43</sup> The inventory contains 21 self-report items which individuals complete using multiple choice response formats. Improvements in severity of depression accompanying study indicate correlation between Vitamin D and depression. | 3 (15%) |
| Depression Diagnostic Criteria (DSM-5)<sup>44</sup> | Depression indicated by changes in appetite, weight, fatigue, sleep and emotional well-being. | If the presence of depression disappears, there is a likely causation between Vitamin D and improved mental health. A more general measurement of depression. Indicates presence of depression, no matter the severity. | 2 (10%) |
| Depression Anxiety Stress Scales (DASS-21)<sup>98</sup> | The DASS-21 is a 21-item self-report instrument assessing depression, anxiety, and stress over the past week. Each item is scored from 0 to 3, and subscale scores (including depression) are summed and multiplied by two to obtain final scores. Severity is categorized into normal, mild, moderate, severe, and extremely severe ranges. | The DASS-21 is a validated tool that captures multiple dimensions of psychological distress, including depressive symptoms. Its multidimensional structure allows for comprehensive assessment of mental health while maintaining reliability in both clinical and research settings. | 2 (10%) |
| Patient Health Questionnaire (PHQ-9)<sup>43,45</sup> | A PHQ-9 score total of 0-4 points equals "normal" or minimal depression. Scoring between 5-9 points indicates mild depression, 10-14 points indicates moderate depression, 15-19 points indicates moderately severe depression, and 20 or more points indicates severe depression. | PHQ-9 is a short screening measure used in medical and community settings to assess depression. If one's PHQ-9 value improves, there is likely a correlation between Vitamin D and improved depressive symptoms. | 5 (25%) |
| Diagnostic Interview Schedule (DIS)<sup>46</sup> | is a questionnaire used to assess symptoms of mental disorders such as depression. | DIS is developed by the National Institute of Mental Health and the detailed description of the questionnaire is described in Plan and Operation of the NHANES III and it is used to make diagnoses based on the third edition of the Diagnostic and Statistical Manual of Mental Disorders (DSM III). | 1 (5%) |
| Hospital Anxiety and Depression Scale (HADS)<sup>99</sup> | The Hospital Anxiety and Depression Scale (HADS) is a 14-item self-report instrument designed to assess symptoms of anxiety and depression in medical settings. It consists of two subscales: HADS-A (anxiety) and HADS-D (depression), each containing 7 items scored from 0 to 3. Subscale scores range from 0 to 21. Standard cutoffs are: 0–7 (normal), 8–10 (borderline abnormal), and 11–21 (abnormal), indicating clinically relevant symptoms. | The HADS is a widely validated instrument developed for use in hospital and clinical populations, specifically designed to minimize the influence of somatic symptoms that may overlap with physical illness. This makes it particularly appropriate for studies involving medical conditions, as it focuses on the psychological aspects of depression and anxiety. Its reliability, validity, and ease of administration support its use in both clinical and research settings. | 1 (5%) |
| Children's Depression Inventory (CDI)<sup>47</sup> | is a questionnaire used to assess symptoms of mental disorders such as depression. The CDI scores symptoms of depression on five subscales that consist of negative mood, interpersonal problems, ineffectiveness, anhedonia, and negative self-esteem with 27 items scored on a 3-point scale<br>Score 0: absence of the symptom<br>Score 1: moderate symptom<br>Score 2: severe symptom | The CDI is specifically validated for pediatric populations and is widely used to assess depressive symptoms in children and adolescents, making it appropriate for studies involving younger participants. | 1 (5%) |
| Ninth Revision of the International Classification of Diseases, Clinical Modification (ICD-9-CM)<sup>100</sup> | The International Classification of Diseases, Ninth Revision, Clinical Modification (ICD-9-CM) is a standardized diagnostic coding system used to classify diseases and health conditions in administrative and clinical records. Depression is identified using specific diagnostic codes (e.g., 296.xx for mood disorders, 311 for depressive disorder not otherwise specified), based on clinician diagnosis documented in medical records. Outcomes are typically recorded as categorical variables (presence vs absence of depression diagnosis). | ICD-9-CM codes are widely used in epidemiological and health services research to identify clinical diagnoses from large administrative datasets. Their standardized structure allows for consistent classification across healthcare systems and enables large-scale population-based analyses. However, as diagnoses rely on clinical documentation and coding practices, they may differ from symptom-based screening tools and can contribute to variability across studies. | 1 (5%) |
| Center for Epidemiologic Studies Depression (CES-D)<sup>48</sup> | Respond options range from 0 to 3 for each item (0 = Rarely or None of the Time, 1 = Some or Little of the Time, 2 = Moderately or Much of the time, 3 = Most or Almost All the Time). Scores range from 0 to 60, with high scores indicating greater depressive symptoms. | CES-D is a 20-item brief multi-perspective assessment of depressive symptoms. CES-D asks individuals to rate how often the past week they experienced symptoms associated with depression, such as restless sleep, poor appetite, and feeling lonely. | 4 (20%) |

**Appendix 6:** Assessment of methodological quality using the Newcastle Ottawa Scale.

| First author, Year | Selection<br>(0-4) cohort<br>(0-5) cross | Comparability<br>(0-2) cohort<br>(0-1) cross | Outcome<br>(0-3) cohort<br>(0-3) cross | Quality (Good, Fair, Poor)* |
| --- | --- | --- | --- | --- |
| Al-Sabah, 2022 | 4 | 1 | 2 | Good |
| Anuroj, 2022 | 3 | 1 | 3 | Good |
| Bigman, 2022 | 3 | 2 | 2 | Good |
| Black, 2014 | 5 | 1 | 3 | Good |
| Callegari, 2017 | 3 | 2 | 2 | Good |
| Ganji, 2010 | 3 | 1 | 3 | Good |
| Kamalzadeh, 2021 | 4 | 1 | 3 | Good |
| Kerr, 2015 | 2 | 1 | 3 | Good |
| Kim, 2020 | 5 | 1 | 2 | Good |
| Kwasky, 2012 | 3 | 1 | 2 | Good |
| Kwasky, 2014 | 3 | 1 | 2 | Good |
| Lasaite, 2011 | 3 | 1 | 3 | Good |
| Ma, 2023 | 4 | 1 | 3 | Good |
| Polak, 2014 | 3 | 1 | 2 | Good |
| Shaad, 2019 | 4 | 1 | 3 | Good |
| Satyanarayana, 2023 | 4 | 1 | 2 | Good |
| Tomlinson, 2021 | 3 | 1 | 3 | Good |

\*Number of stars given using the Newcastle-Ottawa Scale (NOS):
**Good Quality:** 3-4 stars in the selection domain and 1-2 stars in the comparability domain and 2-3 stars in the outcome domain.
**Fair Quality:** 2 stars in the selection domain and 1-2 stars in the comparability domain and 2-3 stars in the outcome domain.
**Poor Quality:** 0-1 star in the selection domain or 0 stars in the comparability domain or 0-1 star in the outcome domain.

**Appendix 7:** Systematic Review Protocol Modifications.

| Date | Modification | Justification |
| --- | --- | --- |
| February 03, 2023 | Limited to studies in English | To avoid any bias that can happen during translation |
| September 30, 2023 | Added to inclusion criteria table:<br>1. Randomized control trial<br>2. Case control studies<br>3. Case control studies<br>4. Case studies | See **Appendix 1** study design section |
| March 28, 2026 | Conduct additional scientificity analyses to explore source of heterogeneity | At the request of reviewers during journal peer-review process and given the substantial heterogeneity observed across pooled analyses, we conducted additional sensitivity analyses to assess the robustness of the findings: (1) leave-one-out analyses, (2) geographic subgroup exploration, and (3) prediction interval analyses. |

**Appendix 8:**
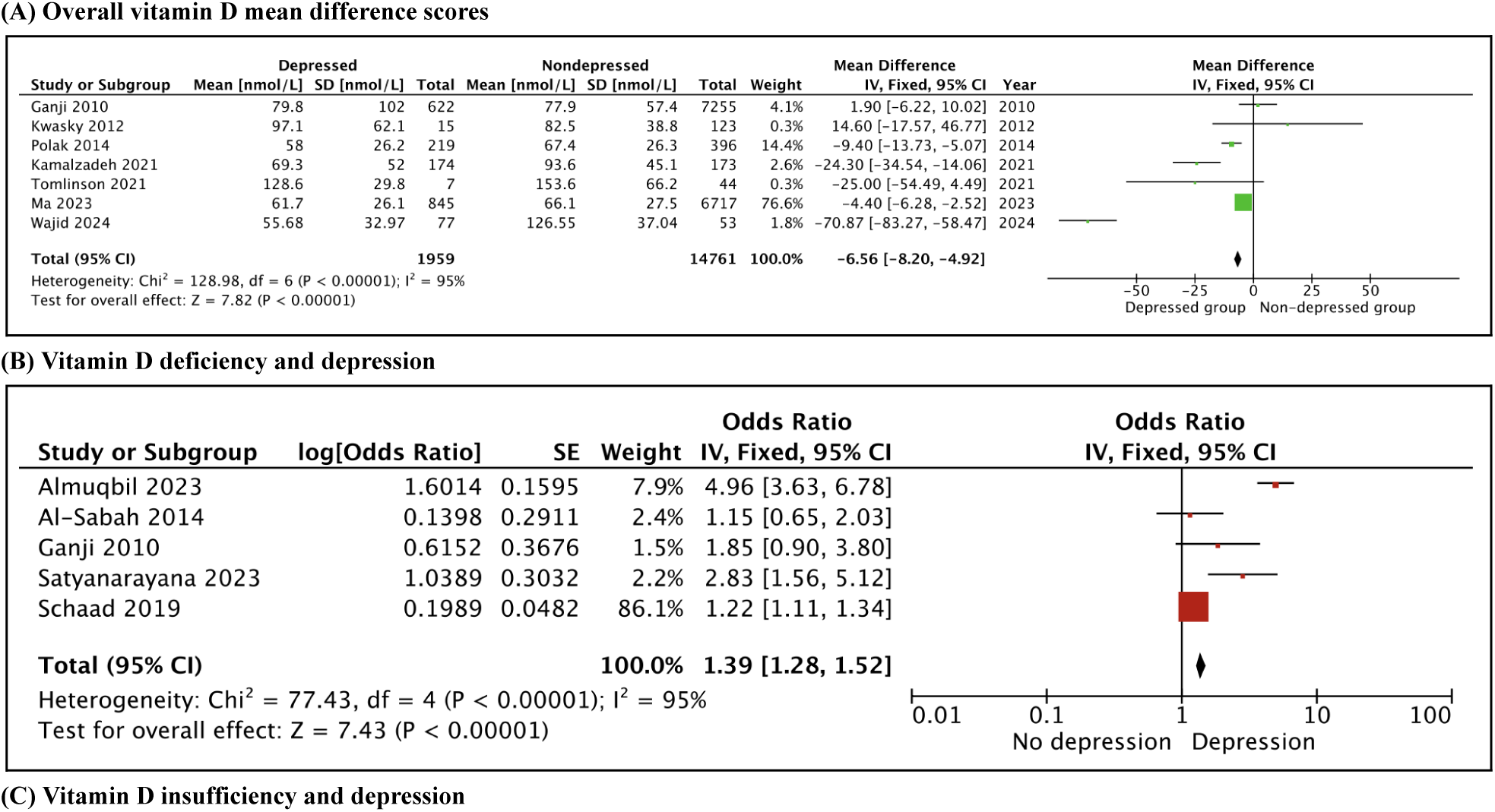
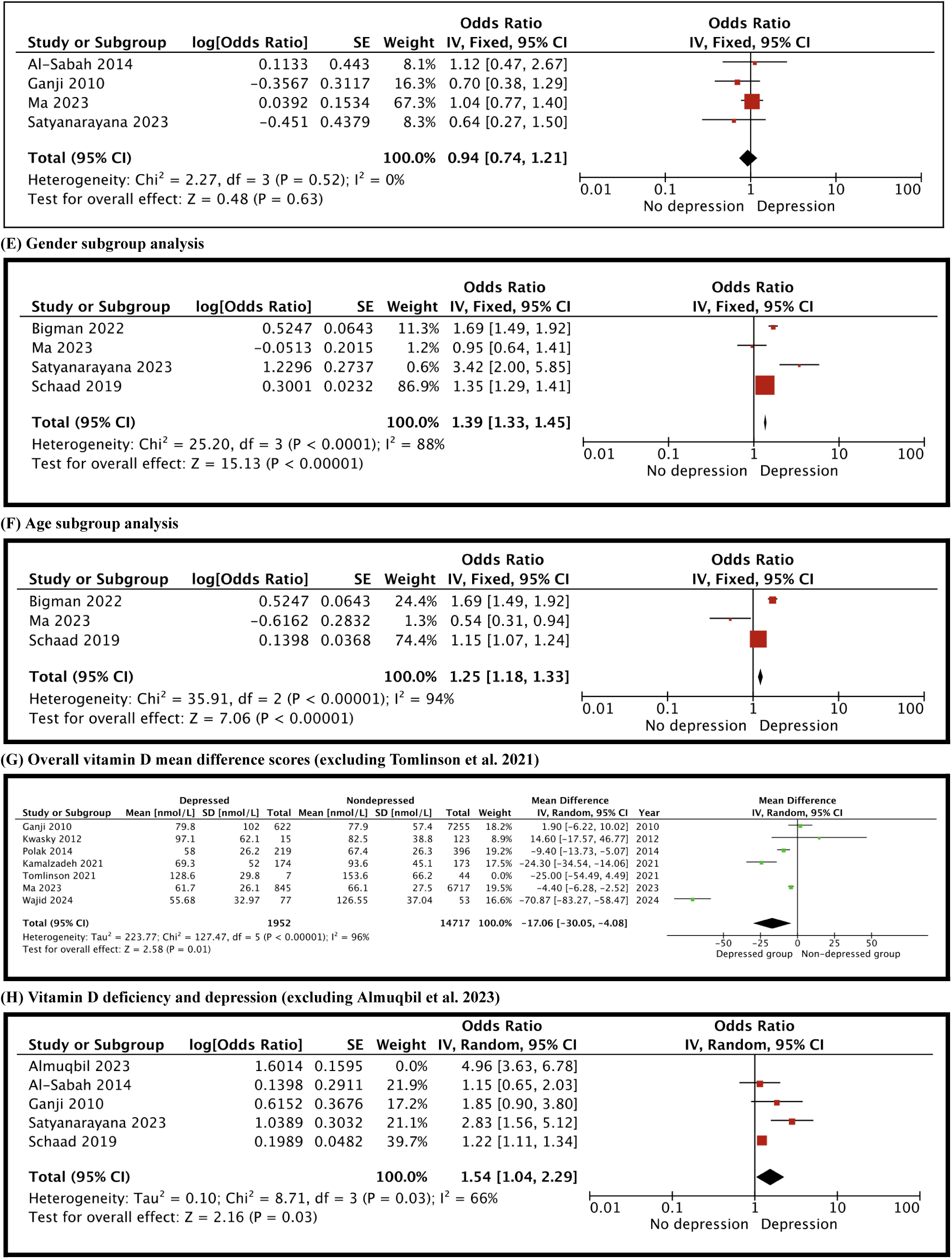
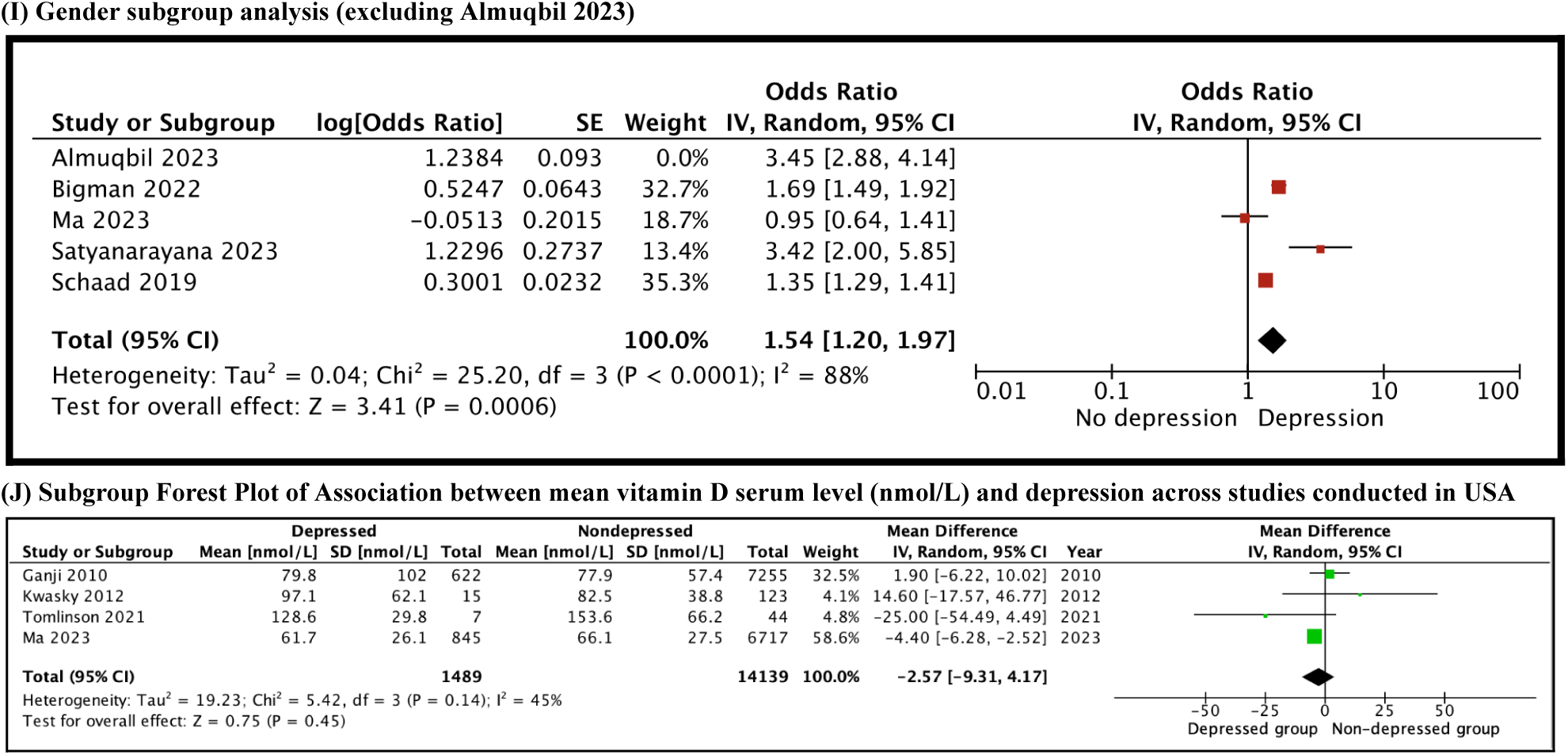
Sensitivity analysis.

**Appendix 9.** Summary of Covariates Included in Adjusted Models Across Included Studies.

| Study (Year) | Sex | Age | BMI | Race-Ethnicity | Other Covariates |
| --- | --- | --- | --- | --- | --- |
| Almuqbil (2023) | √ | - | √ | - | √ |
| Al Sabah (2022) | √ | √ | - | √ | √ |
| Bigman (2022) | √ | √ | - | √ | √ |
| Ganji (2010) | √ | √ | √ | √ | √ |
| Ma (2023) | √ | √ | √ | √ | √ |
| Satyanarayana (2023) | √ | √ | - | - | √ |
| Schaad (2019) | √ | √ | - | - | √ |

**Appendix 10:** PRISMA 2020 Checklist.

| Section and Topic | Item # | Checklist item | Location where item is reported |
| --- | --- | --- | --- |
| **TITLE** |  |  |  |
| Title | 1 | Identify the report as a systematic review. | 1 |
| **ABSTRACT** |  |  |  |
| Abstract | 2 | See the PRISMA 2020 for Abstracts checklist. | 2 |
| **INTRODUCTION** |  |  |  |
| Rationale | 3 | Describe the rationale for the review in the context of existing knowledge. | 3 |
| Objectives | 4 | Provide an explicit statement of the objective(s) or question(s) the review addresses. | 4 |
| **METHODS** |  |  |  |
| Eligibility criteria | 5 | Specify the inclusion and exclusion criteria for the review and how studies were grouped for the syntheses. | 4 |
| Information sources | 6 | Specify all databases, registers, websites, organisations, reference lists and other sources searched or consulted to identify studies. Specify the date when each source was last searched or consulted. | 5 |
| Search strategy | 7 | Present the full search strategies for all databases, registers and websites, including any filters and limits used. | 33-38 |
| Selection process | 8 | Specify the methods used to decide whether a study met the inclusion criteria of the review, including how many reviewers screened each record and each report retrieved, whether they worked independently, and if applicable, details of automation tools used in the process. | 5 |
| Data collection process | 9 | Specify the methods used to collect data from reports, including how many reviewers collected data from each report, whether they worked independently, any processes for obtaining or confirming data from study investigators, and if applicable, details of automation tools used in the process. | 5-6 |
| Data items | 10a | List and define all outcomes for which data were sought. Specify whether all results that were compatible with each outcome domain in each study were sought (e.g. for all measures, time points, analyses), and if not, the methods used to decide which results to collect. | 6, 39-40 |
|  | 10b | List and define all other variables for which data were sought (e.g. participant and intervention characteristics, funding sources). Describe any assumptions made about any missing or unclear information. | 6, 39-40 |
| Study risk of bias assessment | 11 | Specify the methods used to assess risk of bias in the included studies, including details of the tool(s) used, how many reviewers assessed each study and whether they worked independently, and if applicable, details of automation tools used in the process. | 7-8 |
| Effect measures | 12 | Specify for each outcome the effect measure(s) (e.g. risk ratio, mean difference) used in the synthesis or presentation of results. | 6-7 |
| Synthesis methods | 13a | Describe the processes used to decide which studies were eligible for each synthesis (e.g. tabulating the study intervention characteristics and comparing against the planned groups for each synthesis (item #5)). | 7-8 |
|  | 13b | Describe any methods required to prepare the data for presentation or synthesis, such as handling of missing summary statistics, or data conversions. | 4, 7-8 |
|  | 13c | Describe any methods used to tabulate or visually display results of individual studies and syntheses. | 7-8 |
|  | 13d | Describe any methods used to synthesize results and provide a rationale for the choice(s). If meta-analysis was performed, describe the model(s), method(s) to identify the presence and extent of statistical heterogeneity, and software package(s) used. | 6-8 |
|  | 13e | Describe any methods used to explore possible causes of heterogeneity among study results (e.g. subgroup analysis, meta-regression). | 7-8 |
|  | 13f | Describe any sensitivity analyses conducted to assess robustness of the synthesized results. | 7-8 |
| Reporting bias assessment | 14 | Describe any methods used to assess risk of bias due to missing results in a synthesis (arising from reporting biases). | 8 |
| Certainty assessment | 15 | Describe any methods used to assess certainty (or confidence) in the body of evidence for an outcome. | 8 |
| **RESULTS** |  |  |  |
| Study selection | 16a | Describe the results of the search and selection process, from the number of records identified in the search to the number of studies included in the review, ideally using a flow diagram. | 8 |
|  | 16b | Cite studies that might appear to meet the inclusion criteria, but which were excluded, and explain why they were excluded. | 8 |
| Study characteristics | 17 | Cite each included study and present its characteristics. | 8 |
| Risk of bias in studies | 18 | Present assessments of risk of bias for each included study. | 9 |
| Results of individual studies | 19 | For all outcomes, present, for each study: (a) summary statistics for each group (where appropriate) and (b) an effect estimate and its precision (e.g. confidence/credible interval), ideally using structured tables or plots. | 9-11, 27-32 |
| Results of syntheses | 20a | For each synthesis, briefly summarise the characteristics and risk of bias among contributing studies. | 11 |
|  | 20b | Present results of all statistical syntheses conducted. If meta-analysis was done, present for each the summary estimate and its precision (e.g. confidence/credible interval) and measures of statistical heterogeneity. If comparing groups, describe the direction of the effect. | 9-11 |
|  | 20c | Present results of all investigations of possible causes of heterogeneity among study results. | 10 |
|  | 20d | Present results of all sensitivity analyses conducted to assess the robustness of the synthesized results. | 10 |
| Reporting biases | 21 | Present assessments of risk of bias due to missing results (arising from reporting biases) for each synthesis assessed. | 11 |
| Certainty of evidence | 22 | Present assessments of certainty (or confidence) in the body of evidence for each outcome assessed. | 9-11 |
| **DISCUSSION** |  |  |  |
| Discussion | 23a | Provide a general interpretation of the results in the context of other evidence. | 11 |
|  | 23b | Discuss any limitations of the evidence included in the review. | 12 |
|  | 23c | Discuss any limitations of the review processes used. | 12 |
|  | 23d | Discuss implications of the results for practice, policy, and future research. | 12-13 |
| **OTHER INFORMATION** |  |  |  |
| Registration and protocol | 24a | Provide registration information for the review, including register name and registration number, or state that the review was not registered. | 4 |
|  | 24b | Indicate where the review protocol can be accessed, or state that a protocol was not prepared. | 4 |
|  | 24c | Describe and explain any amendments to information provided at registration or in the protocol. | 4 |
| Support | 25 | Describe sources of financial or non-financial support for the review, and the role of the funders or sponsors in the review. | 14 |
| Competing interests | 26 | Declare any competing interests of review authors. | 14 |
| Availability of data, code and other materials | 27 | Report which of the following are publicly available and where they can be found: template data collection forms; data extracted from included studies; data used for all analyses; analytic code; any other materials used in the review. | 14 |

From: Page MJ, McKenzie JE, Bossuyt PM, Boutron I, Hoffmann TC, Mulrow CD, et al. The PRISMA 2020 statement: an updated guideline for reporting systematic reviews. *BMJ* 2021;372:n71. doi: 10.1136/bmj.n71. This work is licensed under CC BY 4.0. To view a copy of this license, visit https://creativecommons.org/licenses/by/4.0/

